# Effects of single plant-based vs. animal-based meals on satiety and mood in real-world smartphone-embedded studies

**DOI:** 10.1101/2021.10.24.21265240

**Authors:** Evelyn Medawar, Marie Zedler, Larissa de Biasi, Arno Villringer, A. Veronica Witte

**Author notes:** Corresponding author: Evelyn Medawar, Department of Neurology, Max Planck Institute for Human Cognitive and Brain Sciences, Stephanstr. 1A, 04103 Leipzig, Germany.

## Abstract

Adopting plant-based diets high in fiber may reduce global warming and obesity prevalence. Physiological and psychological determinants of plant-based food intake remain unclear. As fiber has been linked with improved gut-brain signaling, we hypothesized that a single plant-based (vegetarian and vegan) compared to an animal-based (animal flesh) meal, would induce higher satiety, higher mood and less stress. In three large-scale smartphone-based studies, adults (n_all_ = 16,379) ranked satiety and mood before and after meal intake. Meal intake induced satiety and higher mood. Plant-based meal choice did not explain differences in post-meal hunger. Individuals choosing a plant-based meal reported slightly higher mood before and smaller mood increases after the meal compared to those choosing animal-based meals. Protein content marginally mediated post-meal satiety, while gender and taste ratings had a strong effect on satiety and mood in general. We could not detect profound effects of plant-based vs. animal-based meals on satiety and mood.

## Introduction

Plant-based diets, high in fiber (i.e., non-digestible carbohydrates) stemming from grain products, bread, potatoes, vegetables, legumes and fruits, are linked with planetary ^1,2^ and human health ^3,4^. An extensive meta-analysis showed that higher intake of food items rich in dietary fiber, such as whole-grain products, mediates the benefits of healthy lifestyles on non-communicable diseases such as obesity in a dose-response relationship ^5^. Consequently, increasing daily fiber intake from today’s ∼15 g to 50 g on a global scale, but in particular in Westernized diets, has been proposed to lead to overall extended lifespan and reduced health-care costs ^6^. Eventually, adopting more plant-based diets on a global scale could thus in the long run help to counter ever-increasing rates of obesity and greenhouse gas emissions likewise. Knowledge on the physiological and psychological factors, including hunger and mood related to plant-based meal consumption and food availability and culture, that link to food decision-making remain however surprisingly largely understudied.

On the one hand, several bottom-up physiological mechanisms, prompted by ingested nutrients and processed in the brain, have been proposed to mediate acute metabolic effects signaling post-prandial satiety, reward and contentment, but also long-term metabolic (dys-) regulation and (de-) sensitization ^7^. Macronutrient content most likely modulates appetite control, as energy density or protein content of a single meal determines subsequent metabolic activity ^8,9^ and in turn influences future meal intake via gut-brain hormonal signaling ^10^. In a three-group randomised cross-over study, plant-based compared to macronutrient-matched meat-based meal induced greater satiety independent of weight status ^11^. Indeed, repetitive cueing with highly palatable foods led to habituation of reward-associated dopaminergic signaling ^12^. In contrast to high caloric foods, dietary fiber not only moderately improved body weight status independent of energy intake across 62 trials ^13^, but also activates appetite-regulatory pathways via anorectic hormones, thereby ameliorating high-caloric food craving and other psychobiological processes ^14^. Moreover, in a triple-blind RCT, short-chain fatty acid administration, representative of fermentation products of dietary fiber, led to a reduction in psychosocial stress response after one week ^15^. While the timeframe of those putative effects remain unclear ^16^, fiber and polyunsaturated fats may contribute to healthy feeding-related signaling and neuronal survival, for example by improving glucose and insulin metabolism, by balancing energy homeostasis through protecting hypothalamic neurons from inflammation ^17^ or by contributing to a healthy energy-harvesting profile of the gut microbiome ^18^.

On the other hand, top-down psychological mechanisms include for instance (un-) successful self-control and cognitive strategies towards health or moral goals help to make choices aligned with inner beliefs and self-reinforce dietary habits. Those choices are planned not impulsive, and in the case of food decision-making may often - but not always (e.g. in the case of eating disorders) - lead to healthier food choices ^19^. Pre-meal planning has been shown to be influenced by attentional focus at the time of choice ^20^ – a cognitive control mechanism which may be present in plant-based dieters for every meal planning, yet which has not been investigated. In the case of vegetarians and vegans, restrictive eating has been linked with higher risk for depressive symptoms (meta-analysis of n ∼50k participants ^21^). The determinants, the timeframe and the reasons for the potential link between restrictive diets and depressive mood remain unclear and might include social exclusion, isolation or stigma in the long term. Conversely, healthy and adequate nutrient intake, including high fiber intake, has been associated with lower depressive symptoms and anxiety in observational (meta-analysis, ^22^) and interventional studies ^23^, potentially via microbiota-driven modification of gene expression and anti-inflammatory properties ^24^. Studies investigating short-term effects of food intake related to meal composition on mood are missing to our knowledge. Overall, reverse causation for food-mood relationships remains an unsolved issue ^25^ and whether single meals different in fiber content affect mood remains unclear.

Targeting food choice environment such as labelling, reducing portion size, imposing taxes and modifying availability are common tools to modulate food decision-making ^26^. For instance, a recent study in university cafeterias doubled the availability of vegetarian meals, which led to an increase of 41-79% sales of plant-based meals across over 90,000 meals, without a drop in overall sales ^27^. Also, discounting plant-based meals whilst increasing prices of meat-based dishes led to a slight increase in sales of the former ^28^, whilst order of meals in close proximity did not ^29^. Sales and attractiveness increased when product packaging included labels that prompt sensory or contextual experiences, which was found to be less frequent for plant-based products ^30^. While shaping food choice externally seems a promising tool to change dietary intake, perceived physiological and psychological effects linked to such decisions and meal intakes remain unknown.

In sum, plant-based diets high in fiber resemble the current diet of choice for climate reasons, and some, but not all, interventional studies and meta-analyses raise the hypothesis that plant-based diets contribute to better maintenance of gut-brain signaling including satiety regulation and food reward sensitivity through nutrient-related improvements in metabolic factors. Only few studies however report significant effects of a single plant-based meal on post-prandial satiety and mood, and which factors modify this relation. To address these questions, we aimed to determine satiety and mood before and after a single plant-based (vegetarian, vegan) meal or, as a comparison, animal-based (fish, meat) meal served in university cafeterias providing a broad selection of different meals in the same environment in a demographically homogeneous population. To this end, we designed a series of novel pre-registered smartphone-based studies: firstly, a large-scale study with free meal choice covering most of German university cafeterias, secondly, a study focusing on a smaller, deeply phenotyped sample of omnivorous individuals with free meal choice, and thirdly, a follow-up study of the latter with random allocation of meals.

We pre-registered the following hypotheses:

1. A plant-based (i.e., vegan, vegetarian) meal will lead to better mood, higher satiety and less stress compared to an omnivorous meal.
2. Higher fiber content in the meals will mediate higher mood and satiety and lower stress, whereas higher unrefined sugar and fat content will mediate the opposite.
3. A voluntary decision to eat a plant-based meal (vegetarian or vegan) compared to an omnivorous meal will be more frequently made upon planned (vs. impulsive) decisions.
4. The former potential effects (3) will be masked in participants that followed, and in particular in those that also disliked, a non-voluntary decision of meal category.

We further explored if energy intake (kcal), fluids, environment, dietary habits, social interaction, personality traits and other factors such as age, gender and socioeconomic status modulate the above effects.

## Results

### Sample size, dietary habits and gender distribution (studies 1, 2, 3)

In total 16,135 observations were included in the analysis after data curation for the app study (predominant dietary habits according to self-report: predominantly omnivorous, n = 11,600, predominantly vegetarian, n = 3,456, predominantly vegan, n = 911), as well as 173 in sub-study two and 71 in sub-study three (all omnivorous, with self-reported mean weekly meat intake of about 480 g, representative of a German, Western-style diet with regular meat intake). Reported meals were 47-61% animal-based across studies. Two of the most frequent meals were pasta and currywurst, for study design and exemplary cafeteria meals, see **Figure 1**. Sample size dropped for exploratory data analysis due to missing data (**Supplementary Figure 1**).

**Figure 1:**
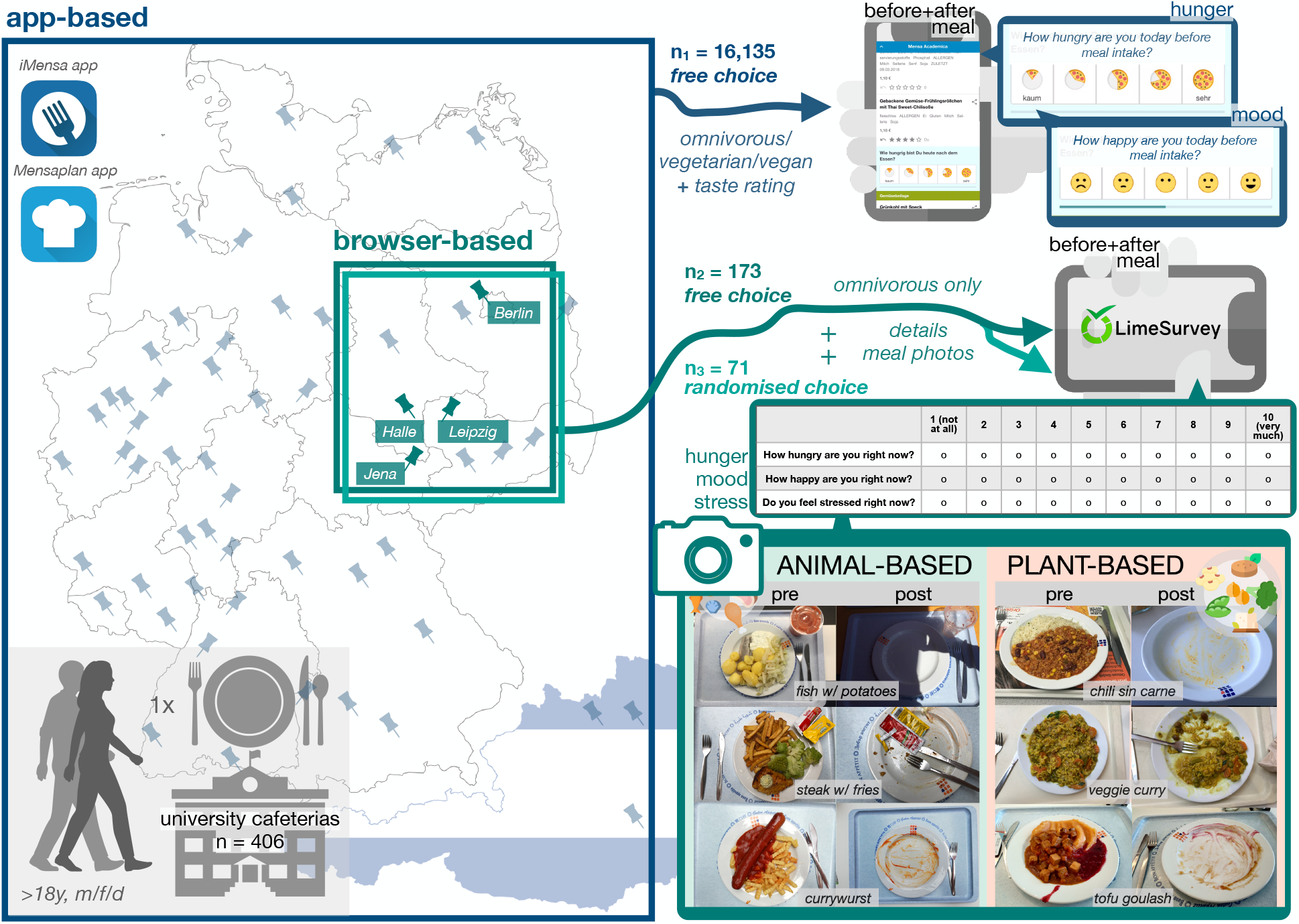
Study design: Adults (m, f, d) consuming a single meal in university cafeterias rated hunger and mood (and stress) using Likert scales before and after that meal with smartphones or mobile devices, in an app study (n_1_ = 16,135, 5-point scales, in dark blue) or in a more detailed web-based survey (omnivorous diet habits only, 10-point scales, n_2_ = 173: free choice (in petrol), n_3_ = 71: randomised choice (in turquoise)). Photos show exemplary pre- and post-meal plates for both meal categories (in petrol). Screenshots taken from iMensa app directly and meal photos provided by study participants (available at https://osf.io/mqc5d/). Maps for Germany and Austria made by Vemaps.com. Copyright for the app icons “iMensa” and “Mensaplan” has been granted by Aimpulse Intelligent Systems GmbH and is under the GNU General Public License for the icon for “LimeSurvey” (https://www.limesurvey.org/, LimeSurvey GmbH, Hamburg, Germany). Images from Flaticon.com (DinosoftLabs and Freepik) and from Apple Keynote were used.

For details about demographic and other parameters see **Figure 2** and **Table 1**. Briefly, in the app study omnivorous dieters chose an animal-based meal more frequently (69%) and vegetarians and vegans mostly chose plant-based meals (84%) (X² (2) = 3679.3, p < 2.2×10^−16^). Note, that a substantial proportion of those reporting to eat predominantly vegan/vegetarian chose an animal-based meal (16%).

**Figure 2:**
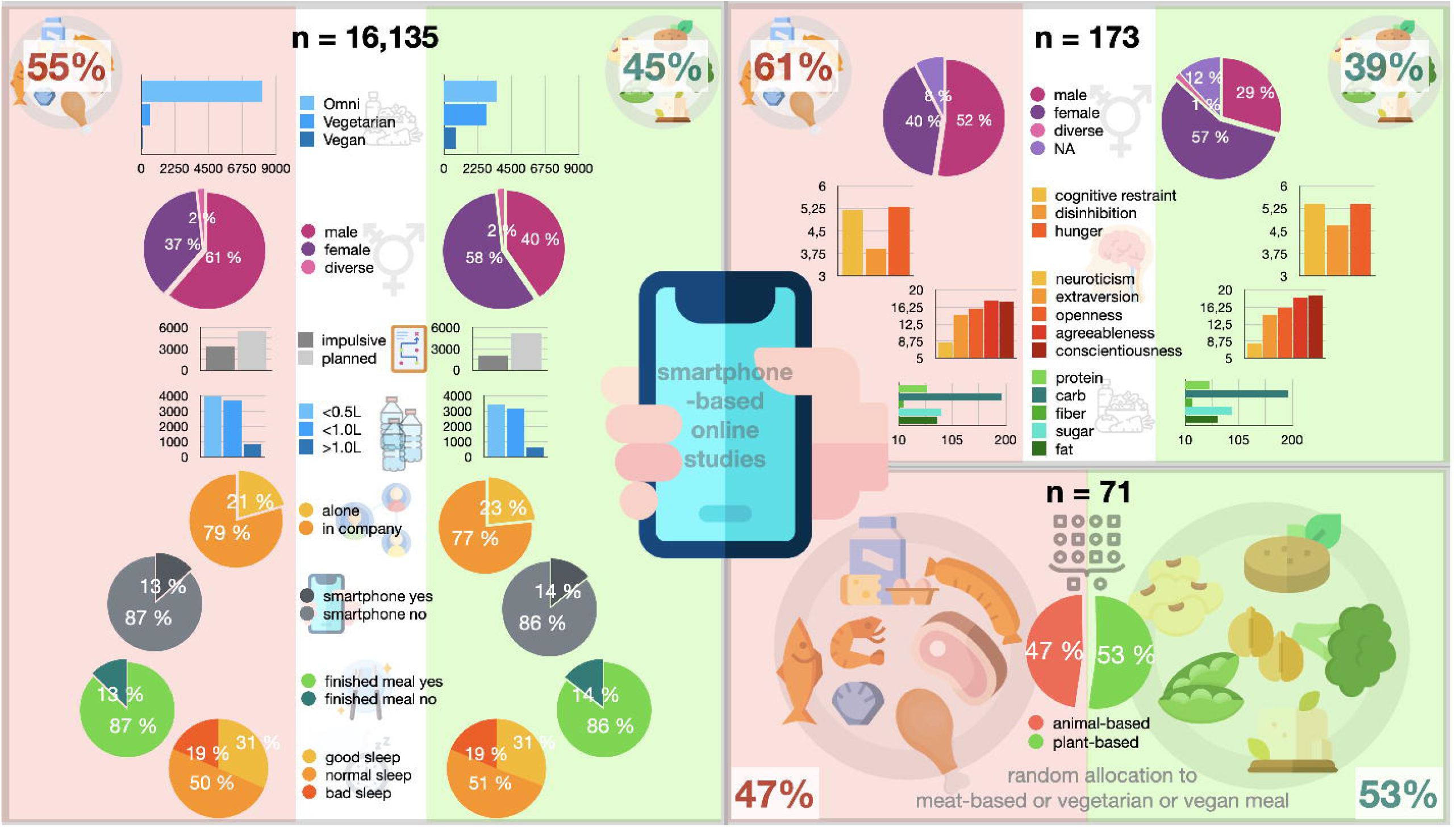
Demographic factors and study sample descriptives of sub-studies 1, 2 and 3. On average, women were more likely to choose a plant-based meal (app study: X² = 731.4, p < 10^−10^, female = 58%, diverse = 45.3%, male = 37.3%; sub-study 2: female = 65%, diverse = 2%, male = 33%; sub-study 3: allocation was randomised). Both female and diverse gender reported more than twice as often predominantly vegan/vegetarian dietary habits compared to male in the app study (X² = 1266, p < 10^−10^, female = 40.4%, diverse = 54.8%, male = 16.5%). In sub-study 2, neither body mass index (BMI), income or general well-being differed across meal categories (p_all_ > 0.26). Omnivorous dieters choosing a plant-based meal were characterized by significantly higher habitual fiber intake (22 ± 11 vs. 18 ± 9 g fiber / day, p < 0.05) and marginally higher scores for cognitive restraint and hunger in the TFEQ subscales (p < 0.05) compared to those choosing animal-based meals (**Table 1**). No differences in personality traits or general well-being were found. Respective differences were not significant for the randomised allocation samples in sub-study 3. Images from Flaticon.com were used: Freepik (smartphone, diet, water, network, eat, clock, broccoli, tofu, peas, chickpea, soy, vegan, sausage, chop, food), Smashicons (brain), DinosoftLabs (chicken leg), surgan (seafood).

### Pre-registered main analysis

#### Effect of meal category on hunger, mood and stress (studies 1, 2, 3)

In the app-study, in contrast to our hypothesis, there was no significant interaction effect of meal category and timepoint for hunger (p = 0.2, **Figure 3A, Table 2**). While individuals choosing a plant-based meal reported lower hunger on average (main effect, ∼ -1/10th points, b = - 0.10, t = - 6.8, model comparison p < .001) independent of timepoint, average hunger dropped around 2 points on the 5-point Likert scale in both conditions (main effect, b = -1.99, t = -179.4, p < 0.001). Considering mood, contrary to our hypothesis, post-meal mood increased ∼ 2/5th points less for individuals choosing a plant-based meal compared to an animal-based meal (interaction effect, post-meal*plant-based: b = -0.06, t = -3.6, model comparison p < .001, **Figure 3B**). In parallel, exploratively, mood was slightly higher in individuals eating a plant-based meal before the meal (animal-based: 3.48±1 points vs. plant-based: 3.52±1 points) and increased in all individuals after the meal (animal-based: +0.26 points, plant-based: +0.20 points). For sub-studies 2 and 3 no significant interaction effects on hunger or mood were observed. There was no significant main or interaction effects for stress. Average values and valence of betas of the effects of interest appeared similar to the app study (for comparison of mean values see **Figure 3C-D, Table 2**).

**Figure 3:**
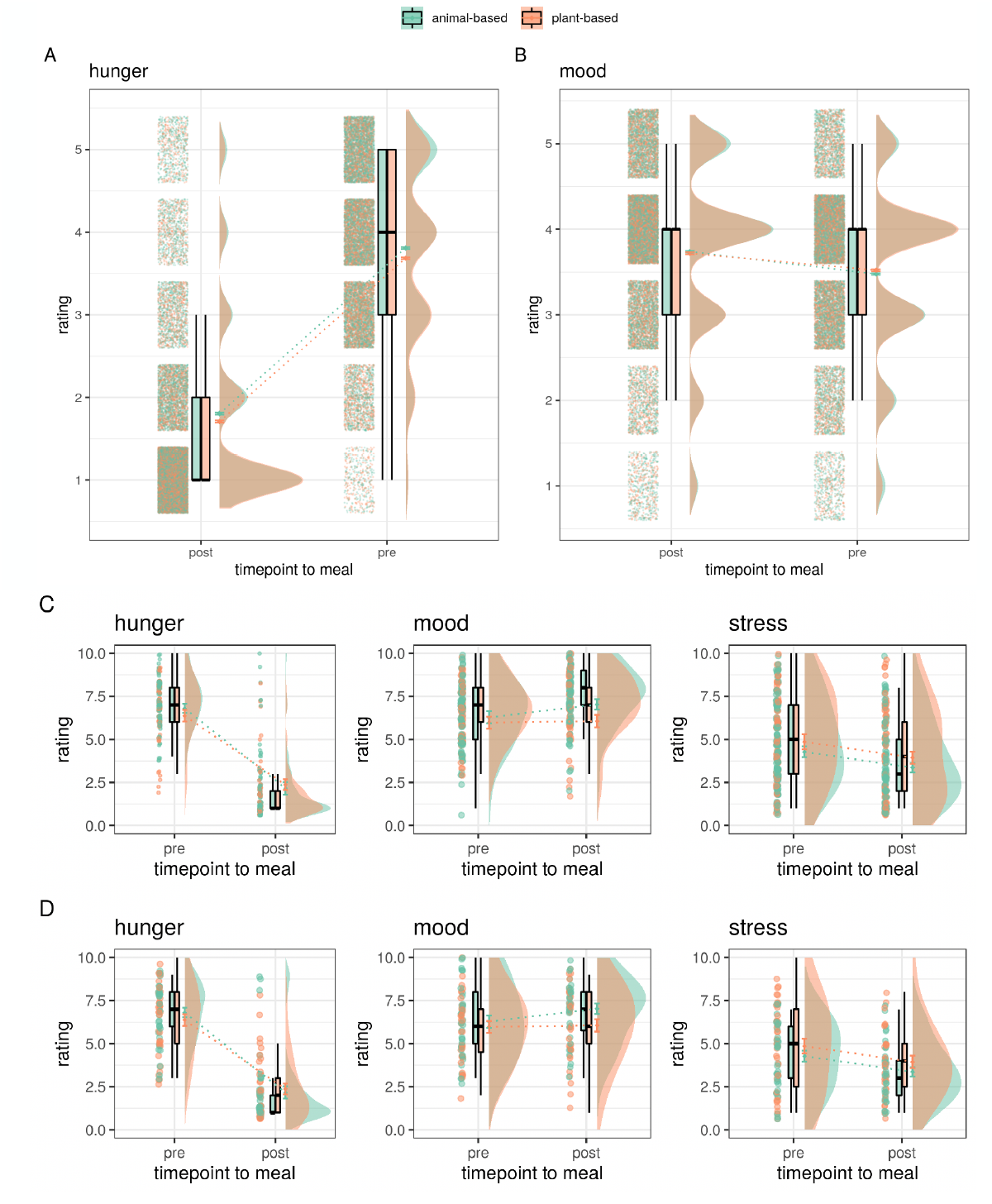
Mean ratings of pre- and post-meal meal category for hunger, mood/contentment and stress. App-study ratings (n = 16135) depicted in A) hunger B) mood, C) sub-study 2 with free meal choice (n=173) and D) sub-study 3 with randomised meal allocation (n=78). Median depicted as line, mean as dot, outliers (according to interquartile rule) as triangles.

### Pre-registered secondary analysis

To further investigate underlying physiological and psychological factors explaining the effects on hunger and mood overall and by meal category, we extended the main analysis and included macronutrient composition available in a subset of datapoints (pre-registered). Also, we investigated if random allocation to a meal, suppressing free choice, would mask any effects. We further explored if taste ratings influenced the main analysis and additionally subdivided the dataset by gender and by dietary adherence (in the app study only; exploratory analysis).

#### Nutrient composition and satiety (study 1)

Macronutrients between meal categories were not significantly different for energy content and saturated fats, but significantly different for carbohydrates, sugar, fat and protein (all Wilcoxon p < 0.01, n_max_ = 1262, **Figure 4, Supplementary Table 1**), with higher carbohydrates, higher sugar, lower fat and lower protein for plant-based meals. The amount of protein, which was approximately one third lower in plant-based meals compared to animal-based meals (**Figure 4F**), had a small effect on post-meal satiety, i.e. that higher protein content led to higher satiety (b = -0.01, t = -3.1, p = 0.002), which was not significantly different between meal categories (interaction effect, p < 0.89). No further significant interaction effects of nutrients were found for the models on hunger and mood.

**Figure 4:**
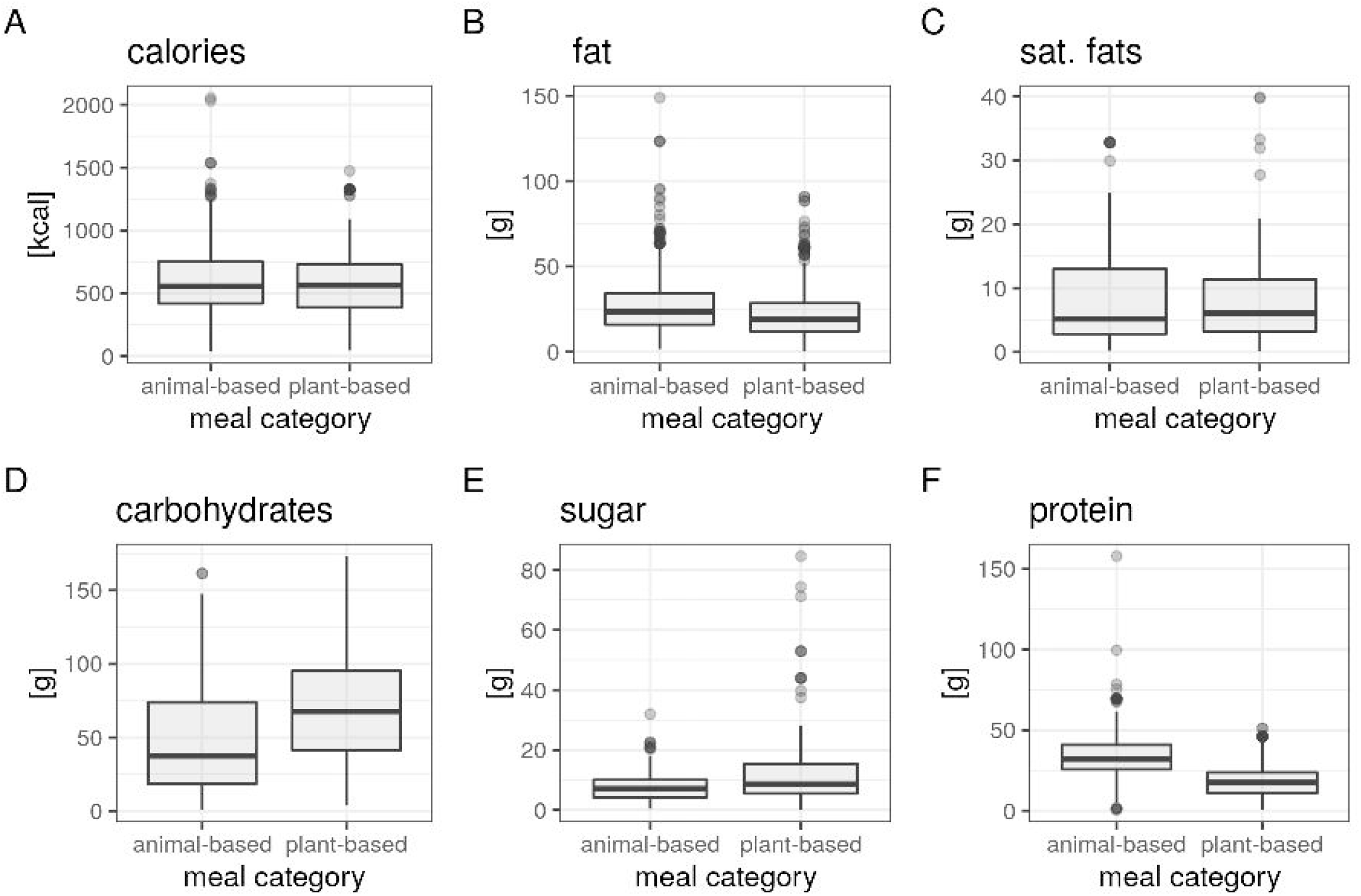
Nutrient composition between meal categories in a subsample of the app-study (n_max_ = 1262, data from app study). Macronutrient information per portion size for A) calories in kcal B) fat in grams C) saturated fats in grams D) carbohydrates in grams E) sugar in grams and F) protein in grams. Median depicted as line, outliers (according to interquartile rule) as dots.

Analyses for fiber content as pre-registered could not performed, due to missing information on fiber content in all cafeterias. We therefore explored differences in meal composition / food components qualitatively in frequency plots of meal components and quantitatively by running models on carbohydrate quality, which is a proxy for fiber content ^31^ (i.e. whole-grain vs. white flour meals). Overall, description of plant-based meals showed higher frequency of vegetables and salads, as well as other food items with high amounts of fiber, like whole grain, lentils or sweet potatoes, compared to animal-based meals (**Supplementary Figure 5**). Further, when comparing identical meal descriptions (most common meal was “spaghetti bolognese”) between white flour (n = 416 meals (animal-based: 317, plant-based=99)) vs. whole-grain flour (n = 26 meals (animal-based: 2, plant-based=24))), we found no significant differences between kcal and protein content (both p > 0.54), and assumed higher fiber content for whole-grain meals. Accordingly, linear mixed models (LMMs) showed that hunger was lower (b = -0.30, t = -1.7, p < 0.2 ×10^−15^) and mood was higher (b = 0.15, t = 0.8, p < 0.8 ×10^−7^) independent of timepoint for spaghetti bolognese made of whole-grain flour compared to white flour. Post-meal ratings did not differ for hunger (b = -0.08, t = -0.28, p = 0.78), mood was significantly increased after whole-grain meals (b = 0.41, t = 1.98 p < 0.05) (**Figure 5**). Comparisons between meal category were not possible due to limited group size for carbohydrate quality per meal category.

**Figure 5:**
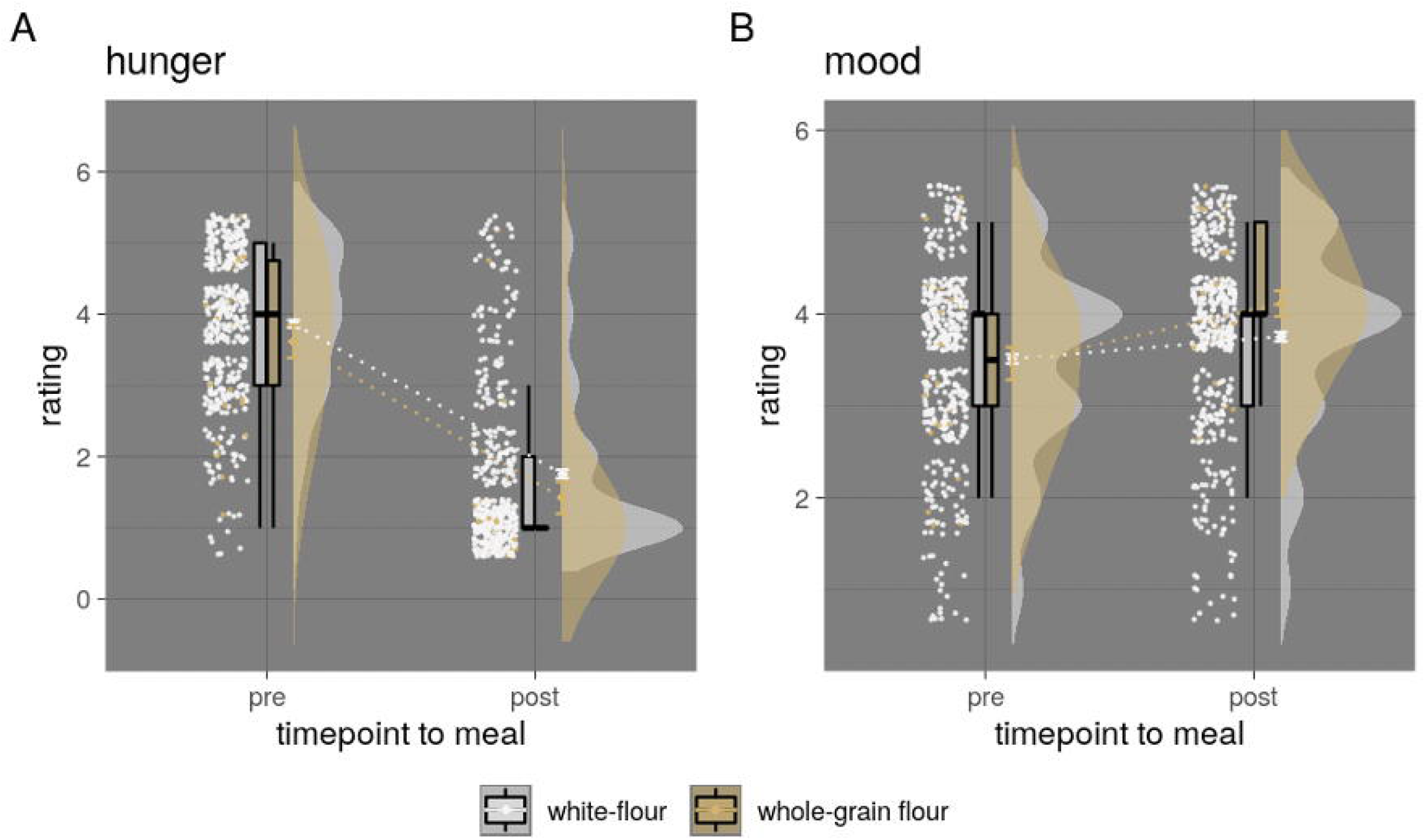
Mean ratings of pre- and post-meal for hunger and mood by white flour vs. whole-grain flour bolognese meals (app-study only (n = 442) depicted in A) hunger B) mood. Median depicted as line, mean as dot, outliers (according to interquartile rule) as triangles.

#### Effects of pre-meal planning in meal choice (study 1)

In the app study, choosing a plant-based meal was 10% more often reported to be a planned decision, compared to that of an animal-based meal choice (X² = 174, p < 2.2×10^−10^, planned plant-based = 72%, planned animal-based = 62%, **Table 1**). Moreover, we observed an interaction effect between the feeling of planned decision-making at meal choice on higher hunger (b = 0.04, t = 2.8, p = 0.006) and higher mood (b = 0.13, t = 8.9, p < 2.2×10^−16^) after the meal. The interaction effect of a planned decision on higher hunger at post-meal was significantly lower for plant-based meal choices (decision*post-meal*plant-based: b = -0.04, t = - 0.9, p < 2×10^−5^), as well as less pronounced for lower mood (decision*post-meal*plant-based: b = -0.02, t = -0.4, p < 5×10^−6^) compared to animal-based meals.

#### Contentment when randomised to meal (study 3)

In sub-study three meal choice was randomly allocated and therefore not a confounding variable. Meal category was randomly assigned after filling out pre-questionnaires right before the planned meal. Contentment about randomization before the meal was not significantly different for animal-based versus vegetarian or vegan meals (n = 71, X^2^ = 9.6, df = 9, p = .27, **Figure 6A**). Overall post-meal contentment after randomization was met more frequently for the animal-based condition (n = 71, X^2^ = 6.1, df = 2, p = .04866, **Figure 6B**). Those who reported to be content with the randomization, did not differ between reasons for it (“chosen anyway”, “preferred something else, but I liked it”, “generally no preference”, X^2^ = 0.03, df = 2, p = .98, **Figure 6C**), compared to those who reported discontent (n = 8 “preferred vegan”, n = 9 “preferred meat-based”, X^2^ = 13.2, df = 1, p < 0.001, **Figure 6D**). However, randomization contentment pre- and post-meal as a main factor did not explain a significant amount of variance with regard to satiety or mood (all betas < 0.28, model comparisons all p > 0.52).

**Figure 6:**
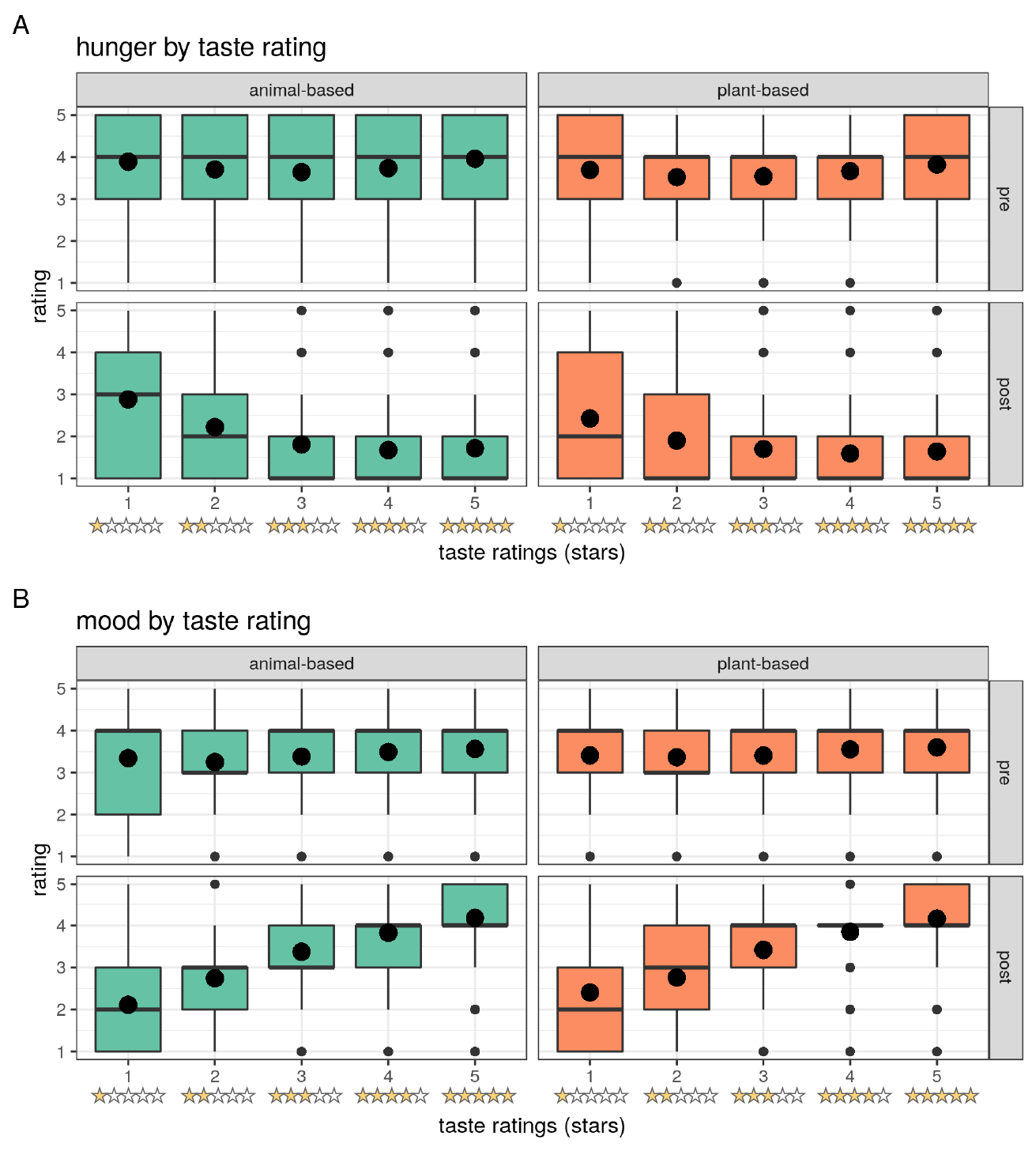
Post-randomization contentment and post-meal contentment and reasons for the latter (sub-study 2, n = 71). A) Pre-meal allocation contentment rating (1-10 Likert scale) by meal category. Mean depicted as dot. B) Post-meal allocation contentment in categorical answers depicted as frequency by meal category. C) Reasons for post-meal contentment in categorical answers depicted as frequency by meal category. D) Reasons for post-meal discontentment in categorical answers depicted as frequency by meal category.

Further, we tested the effect of assigned meal and subsequent liking as preregistered by defining three categories, namely voluntary (n = 174) vs. non-voluntary-liked (n = 49) vs. non-voluntary-disliked (n = 17) of meal category across both sub-studies two and three. Mode of choice did not explain any significant variance on hunger, mood or stress levels across time on meal choice (preregistered model: all p > 0.14). When looking at interaction of timepoint and mode of choice and meal category, post-meal hunger was higher for non-voluntary-liked (b = 1.4, t = 1.8) and lower for non-voluntary-disliked (b = -1.6, t = -1.4) compared to voluntary choice for plant-based meals (model comparison p < 0.004). Post-meal effects of mode of choice for plant-based meals showed higher mood for non-voluntary-liked (b = 0.34, t = 0.5) compared to non-voluntary disliked (b = -0.7, t = -0.7) (model comparison p = 0.028). There was no interaction effect of mode of choice on stress levels post-meal for plant-based meals (p > 0.9).

### Exploratory analysis

#### Effects of taste on hunger and mood independent of meal category (study 1)

Post-meal taste ratings provided on a Likert scale from one to five stars were available in the app study only and showed significant differences between meal categories, with animal-based meals being rated higher in taste more frequently (mean ± SD: animal-based: 3.86 ± 1.23; plant-based: 3.77 ± 1.27, Kruskal-Wallis χ^2^(1) = 21.8, p < .001, scale from one to five stars; zero stars for retracted rating, **Figure 7**).

**Figure 7:**
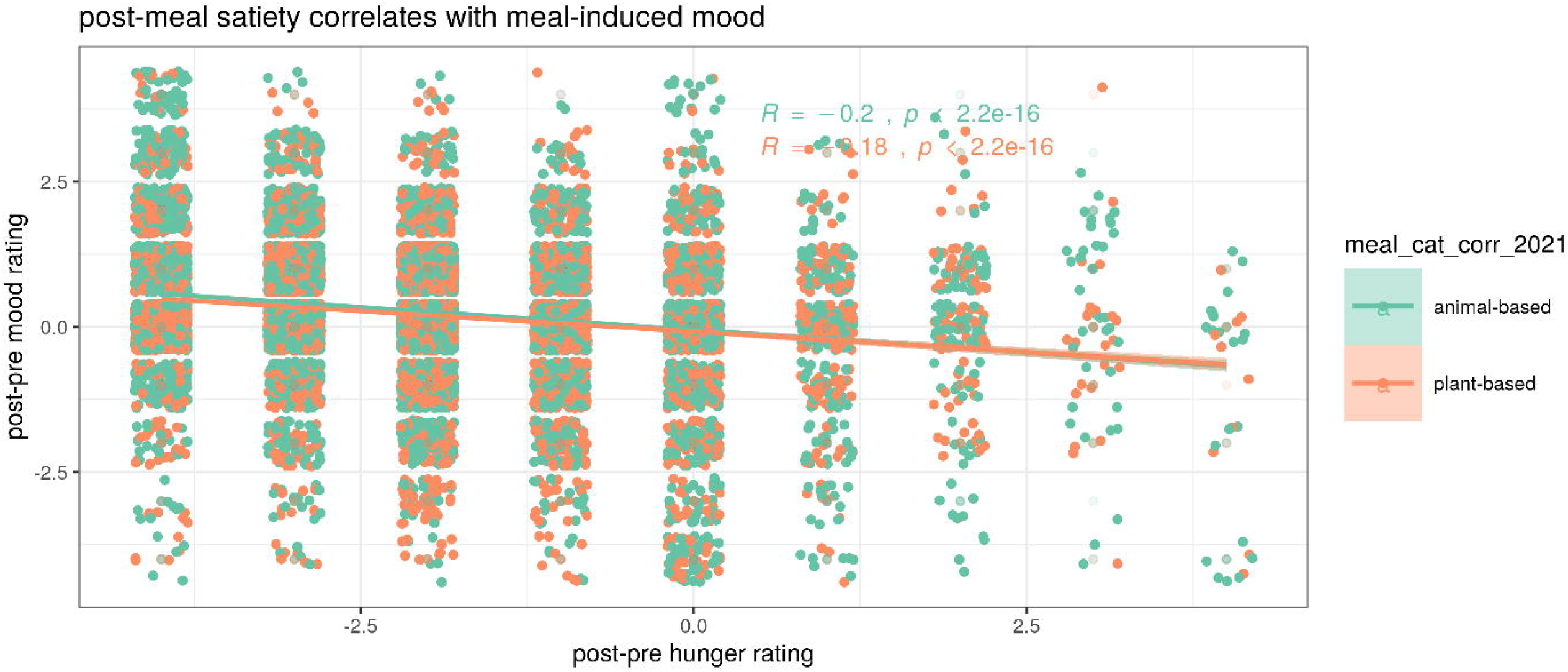
Effects of taste ratings on hunger and mood by star rating provided (app study only). A) Hunger ratings by taste rating from 1 to 5 stars B) Mood ratings by taste rating from 1 to 5 stars. Median depicted as line, mean as dot, outliers (according to interquartile rule) as small dots.

According to LMMs, better taste was related to higher satiety and higher mood on average, with a somewhat linear relationship from lowest to highest taste category, in particular for post-meal mood ratings (**Figure 7, Supplementary Table 2**). Note that only 4.9% of individuals reported lowest taste ratings, and 2.4% retracted the rating (corresponding to zero stars), whereas 35.2% reported maximal ratings across all meals. Overall, low taste ratings increased hunger and high taste ratings lowered hunger considerably (post-meal*1 star: b = 0.6, t = 6.4; post-meal*5 stars: b = -0.5, t = -7.3). Also, low taste ratings had a strong negative effect on post-meal mood (post-meal*1 star: b = -1.3, t = -19.8), whereas high taste ratings moderately elevated post-meal mood (post-meal*5 stars: b = 0.5, t = 8.5). This taste effect on hunger and mood was significantly different between animal-based meals compared to plant-based meals, showing a slightly stronger effect of taste on both hunger and mood after an animal-based meal (**Figure 7**: triple interaction between timepoint, meal category and taste: all betas > |0.01|, all p < 0.01) (**Supplementary Table 2**).

#### Sensitivity analyses for the effects of gender, social interaction, finishing a meal (study 1)

Plant-based meals were more frequently chosen by female and diverse gender, eaten alone and less often finished compared to animal-based meals (**Table 1**). To investigate the potential confounding effects of these factors, we repeated the main models for satiety and mood including these factors in separate models.

Overall, hunger was significantly different for gender, showing lower hunger in female (b = - 0.23, t = -15.6) and higher hunger in diverse (b = 0.21, t = 4.0) compared to male gender (model comparison p < 2.2×10^−16^, **Figure 8A, Supplementary Table 3**). Mood in general was slightly lower in female (b = -0.01, t = -0.9) and drastically lower in diverse gender (b = -0.71, t = -13.8) (model comparison p < 2.2×10^−16^, **Figure 8B, Supplementary Table 3**). When running gender-stratified LMMs (male n = 8291, female n = 7418, diverse n = 279), post-meal hunger after plant-based compared to animal-based meals was similarly higher for male (b = 0.07, t = 2.2, p = 0.03) and female (b = 0.09, t = 3.0, p = 0.003) and not significantly different for diverse (b = - 0.08, t = -0.4, p = 0.7). Models for post-meal mood were significant for both male and female (all p < 0.005), but not for diverse gender (p = 0.08), with similarly lower mood ratings in both males and females (male: b = -0.07, t = -2.8; female: b = -0.1, t = -4.1, diverse: b = 0.35, t = 1.8) after plant-based vs. animal-based meals. Note, that taste ratings were similar across all genders (**Supplementary Figure 7**).

**Figure 8:**
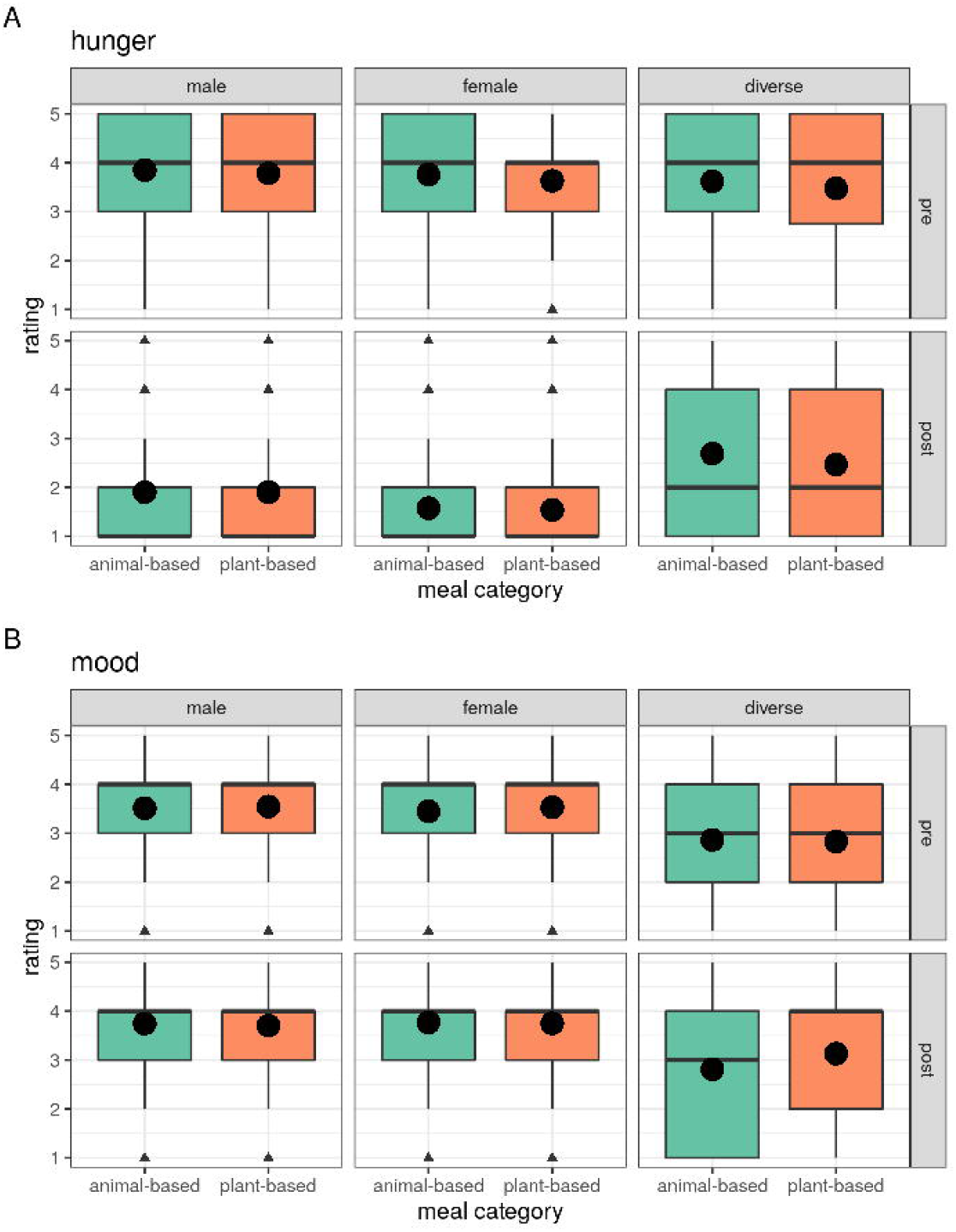
Effects of gender on hunger and mood (app study only, n_male_ = 8291, n_female_ = 7418, n_diverse_ = 279). Median depicted as line, mean as dot, outliers (according to interquartile rule) as triangles.

Eating in company compared to eating alone was related to moderately higher hunger (b = 0.1, t = 6.1) and higher mood (b = 0.1, t = 6.4) (model comparison all p < 2.1×10^−10^). Yet, social company during meal intake did not explain significant variance of post-meal ratings for hunger or mood by meal category (model comparison all p < 0.68).

Finishing a meal was related to higher overall hunger (b = 0.1, t = 5.4) and higher mood (b = 0.4, t = 20.2) (model comparison all p < 7.0×10^−8^). Moreover, finishing a meal also explained slightly higher post-meal hunger ratings (b = 0.05, t = 0.8) but also slightly higher mood (b = 0.08, t = 1.6) for plant-based compared to animal-based meals (model comparison all p < 2.2×10^−16^).

#### Subgroup analysis – dietary habits and time stamps (studies 1, 2, 3)

To account for potential effects of dietary adherence, we repeated the analysis in subgroups for predominantly omnivorous (n = 11,600), predominantly vegetarian (n = 3,456) and predominantly vegan (n = 911) dieters (see **Supplements, Supplementary Table 4, Supplementary Figure 8**). Predominantly omnivorous dieters showed comparable results as the whole-group analysis, i.e. lowered mood and higher hunger after a plant-based meal. Analysis for the predominantly vegetarian group replicated for mood only, not hunger. Contrastingly, predominantly vegans showed higher mood and lower hunger after a plant-based meal. Taste ratings differed by dietary group, such that predominantly omnivores and predominantly vegetarians preferred animal-based meals, yet the opposite was true for predominantly vegans.

Further, we ran sensitivity analyses with restricted time frames around entry time to define a reasonable timeframe around actual food intake (see **Supplements**). Time difference between pre- and post-meal hunger entries in the app study varied quite drastically and was very small for most entries (mean ± SD: 54 ± 139 min, median: 0.5 min), whereas post-pre meal time difference in sub-studies 2+3 inferred from photo time stamps, was rather small with low variance (mean ± SD: 15 ± 7 min, median: 14 min, min: 5 min, max: 40 min), reflecting high compliance with the study design.

#### Correlation of post-meal satiety, mood and stress (studies 1, 2, 3)

Changes in satiety pre- to post-meal correlated with changes in mood pre- to post-meal, i.e. lower meal-induced hunger was associated with higher meal-induced mood, significant for data from the app study (Spearman’s correlations 95%CI p-value; app study: r = -0.19 [-0.21 -0.18] p < .001; sub-study free choice: r = -0.08 [-0.23 0.07] p = 0.3; sub-study randomised allocation: r = 0.13 [-0.10 0.35] p = 0.27, **Figure 9, Supplementary Figure 6**). There was no association between changes in hunger and stress levels (sub-study free choice: r = 0.15 [-0.004 0.288] p = 0.056; sub-study randomised allocation: r = -0.10 [-0.32 0.14] p = 0.43) in the two sub-studies. There was a significant correlation between lower mood and higher stress levels pre- to post-meal in both sub-studies (sub-study free choice: r = -0.34 [-0.46 -0.20] p < 0.001; sub-study randomised allocation: r = -0.39 [-0.57 -0.17] p < 0.001, **Supplementary Figure 6**).

**Figure 9:**
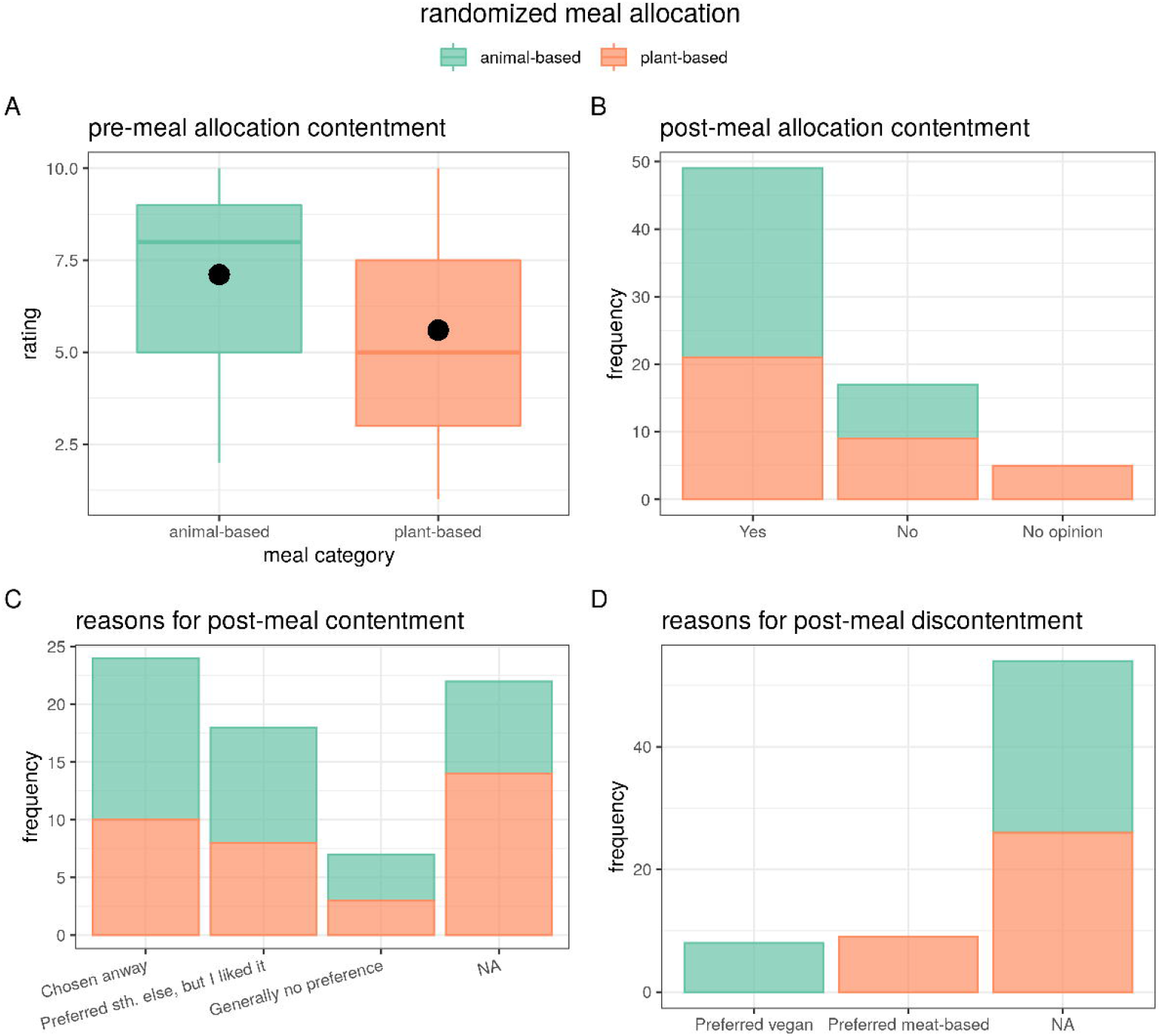
Correlation of post-pre changes between hunger and mood levels plotted by meal category (green: animal-based; orange: plant-based) (app study only). Spearman’s correlation and 95% CI depicted.

## Discussion

In this series of large-scale online surveys of >16,300 individual data entries and three independent data collections in German cafeterias, we provide evidence based on pre-registered analyses that the category of a single meal (i.e. whether plant-based or containing meat/fish) exerts no effect on post-prandial hunger of the individual, and a certain, yet not considerably large influence on mood. Against our hypotheses, there was no significant interaction effect of meal category and timepoint for hunger, but for mood, however contrary to our hypothesis, with smaller post-meal increases in mood for individuals choosing a plant-based meal compared to an animal-based meal. The more deeply phenotyped sub-studies 2 and 3, profoundly smaller in sample size, indicated similar nominal differences between meal categories, yet did not support that those differences should be regarded statistically significant in the main pre-registered analyses. Additionally in exploratory analyses, we found that while participants report higher satiety and improved mood after a meal in general, mood was slightly higher in individuals choosing a plant-based meal before the meal.

Against our hypothesis, that plant-based meals would increase post-meal mood more strongly than animal-based meals, we found the opposite: namely, that post-meal mood was higher after an animal-based meal. Here, we might have neglected certain normative expectations towards the meal in a society that on average consumes meat and fish on a daily basis ^32^, in particular potential reservations against plant-based meals related to be lacking in something or not being hearty enough, which is also in line with in lower taste ratings of plant-based meals. These expectations might have influenced post-meal mood ratings. Yet, for a sub-analysis comparing whole-grain vs. white-flour meals, we found increased post-meal mood for whole-grain meals, supporting the initial hypothesis. Surprisingly, individuals choosing plant-based meals showed higher mood ratings before the meal, which is in contrast to meta-analytical evidence for higher depressive symptoms in long-term vegetarian/vegan dieters ^21^, but in line with improved mood related to pre-/probiotic diets ^22^. Note, that the short timeframe of a single-meal on mood ratings compared to epidemiological evidence of long-term dietary habits could explain the deviating results, and that the effect size of the present study must be considered small. Further, fiber-induced improvements on mood could be particularly effective for vulnerable individuals only, as shown for patients suffering from inflammatory bowel syndrome or major depressive disorder ^23,33^. While we assume that the majority of individuals was healthy in our studies assessing detailed medical history was out of scope in the presented online studies. Besides meal category, taste ratings showed a substantial impact on post-meal mood ratings, with higher mood for more tasteful meals. Importantly, as plant-based meals were ranked less tasty overall, especially by predominantly omnivores and vegetarians, cafeterias should aim to improve tastiness of plant-based meals in order to ensure equal enjoyment and satisfaction despite meal category. When limiting the analysis to identical meal descriptions such as spaghetti bolognese, note that those providing higher fiber content through whole-grain pasta induced higher satiety and higher mood compared to conventional pasta.

We hypothesized that plant-based meals would increase satiety due to a higher amount of fiber in those meals, which we could not evaluate due to missing information on actual fiber content. Further, in a sub-analysis comparing whole-grain vs. white-flour meals, we found no post-meal difference in hunger. This might imply that fiber content has only negligible effects on satiety, and/or that fiber was not different between meal categories on average. In addition, we found that plant-based meals had higher carbohydrate, higher sugar, lower fat and lower protein content compared to animal-based meals in a subset of available datapoints. This is insightful given that plant-based food consists of more carbohydrates, while meat and fish have more protein. Now, in a secondary analysis, we found that protein content contributed to satiety after the meal, arguing that plant-based vs. animal-based meals could relate to lower satiety due to the difference in protein content between meal categories, thereby concealing (potential) effects of higher fiber content on satiety. Indeed, cross-over within-subject studies reported that post-meal satiation did not differ dependent on protein source when adjusted for fiber content ^34^ or in case of protein preload previous to ad libitum food intake ^35^. Moreover, cafeteria meals overall might contain more processed and less balanced nutrient compositions compared to the average plant-based meal investigated in previous literature. Meals containing more plant-derived components are not necessarily healthier, as fast foods and ultra-processed foods, e.g. meat or dairy substitutes, are becoming more prevalent, oftentimes having similar or worse nutrient composition than their animal-based counterparts. Whereas some may have higher sugar and salt content, yet replacements may contribute to improved diet quality by higher fiber and micronutrient levels ^36^. Note that highly processed vegan meals, still four times higher in fiber, were also found to induce higher satiation in a within-subject randomised clinical trial compared to energy-matched meat-containing processed meals ^37^. Another explanation for the lack of strong differences between meal categories might be that meals of both categories were composed of fiber-rich side dishes such as broccoli, thereby “bypassing” the hypothesized disadvantageous effect of animal-based meals on post-meal hunger ratings, potentially leading to comparable fiber content. Notably, participants choosing plant-based meals reported less hunger overall compared to those making animal-based choices. We speculate that this may be due to lower hunger pre-meal choice or due to metabolic adaptations of possibly eating plant-based meals frequently, so that satiety could not be achieved in similar magnitude between meal categories. Future studies could avoid this phenomenon through implementing a more controlled environment with e.g. a 12 h-fasting condition before the meal and thus comparably higher hunger before the meal. However, our large-scale analysis offers the advantage to study effects of a single meal in a real-world context. Thus, while participants choosing a plant-based meal in cafeterias were not more satiated by their current (fiber-rich) meal, they appeared less hungry on average, which might relate to more long-term effects of differences in dietary habits (see below). However, these considerations remain highly speculative given the timeframe of our study design.

Considering biopsychological mechanisms, taste ratings, which were overall slightly higher for animal-based meals, had a strong effect on satiety and mood, highlighting that taste shapes satiety and mood acutely after a plant-based meal and even more so after an animal-based meal. It has been shown that omnivorous individuals expect plant-based alternatives to be less tasty compared to a meat burger ^38^. While impulsive decisions were more frequently made for animal-based meals, such a spontaneous decision further augmented the effect of taste on satiety and mood as shown by out data, whereas in plant-based meal eaters planned decisions had less effect on hunger and mood. Taste have also been shown to influence post-meal satiety ^39,40^, with greater satiation for tasteful meals independent of nutrient composition. Thus, while macronutrients dense in calories such as fat are often described as flavour carriers, and shown to elicit even supra-additive value if paired with carbohydrates on a neuronal level ^41^, low-calorie meals such as salad bowls may induce similarly strong satiety like mac’n’cheese if perceived similarly tasty. However, interoceptive signalling via emotion regulatory processes might determine the properties of any given food further, which may partly explain why some (often high-caloric) foods, such as pizza or burgers, are considered emotionally comforting and rewarding ^42^. Notably, besides the subjective component of taste, there could have been an objective difference in taste by meal category. Meat- and fish-based meals might have been higher in quality due to a longer tradition and knowledge of meal preparation for such ^43^ compared to rather recent developments of vegan and vegetarian cuisine in Germany, that is barely taught in conventional cooking trainings. Moreover, societally meat dishes are viewed as more valuable and might therefore have a higher intrinsic value for omnivorous eaters in meat-heavy culinary traditions ^44^. This may be one of the many reasons why despite wanting to eat more sustainably only some individuals actually succeed in avoiding meat completely, while others continue eating meat occasionally, so called flexitarians ^45^. Considering the university setting, choosing meat-based meals could also be motivated by maximizing food reward linked to expected taste for meaty meals. Although we did not evaluate meal price in this study, oftentimes in cafeterias plant-based meals subsidize animal-based meals leading to equal or even lower prices for meaty meals. Yet, in our analysis socioeconomic status was not a significant predictor of meal choice. Indeed, we also found that hunger and mood changes are not completely independent, but that higher meal-induced satiety correlated with stronger mood improvements.

As hypothesized we found that plant-based meals were more frequently made upon planned decisions compared to impulsive meal decisions. To our knowledge, there are no concrete studies on meal planning in vegetarians and vegans, yet more planning seems expectable, due to constraints in choice from only a subset of meals and meeting pre-defined dietary goals that are in line with their vegetarian identity ^46^.

Further, even though contentment about meal category at time of being informed about randomization was not different between meal groups, post-meal contentment about the allocation differed, with more participants reporting that they would have preferred an animal-based meal when they were dissatisfied with the allocation. Yet, hunger and mood ratings remained unaffected, highlighting that individual preference for category per se did not affect satiety and mood that much. However, when meal category was randomly allocated hunger and mood seemed to be higher when the meal was liked, and both were lower when disliked, showing opposite effects in this study setting compared to the free choice setting. This indicates that a forced choice might enhance the importance of the subjective liking of a certain meal category on hunger and mood, a notion that can inform policy-making when aiming to increase plant-based dieting.

Moreover, gender and whether the individual finished their meal or not could explain a high share of hunger and mood rating differences, whereas social interaction during meal time was not explanatory of post-meal hunger and mood ratings. Previously, evidence for gender differences in eating behaviour ^47,48^ and the influence of others at meal intake ^49,50^ and the influence of finishing or not a meal on reported satiety ^51^ have been shown. Female and diverse gender showed differential effects on hunger and mood ratings related to general meal intake, yet for post-meal ratings for plant-based meals females and males showed similar effects for lowered mood, yet this effect was not present in diverse gender (however note the small sample size for diverse in comparison). Indeed psychological mechanisms of food intake differ by gender, i.e. females score higher on unhealthy eating traits ^48^, show higher prevalence of eating disorders, body and shape concerns ^52^, are more frequently and more strictly on plant-based diets ^47^, follow restrictive diets and weight loss strategies more often, as well as having an increased awareness for health ^53,54^. Only few studies considered gender diverse individuals, yet some evidence supports a heightened risk for eating disorders in sexual and gender diverse students ^55^. Moreover, future studies should consider drink intake of all kinds, including coffee, in the time previous to meal intake. In sum, these findings underline the importance of designing gender-sensitive public health strategies with regard to diet, e.g. tailoring differences in health awareness, mood and emotion regulation and openness to plant-based meals to the individual.

Subgroup analysis suggests that in particular self-reported predominantly omnivorous and predominantly vegetarian dieters are driving the observed interaction effects of meal category on mood but also on hunger in the subsample analysis, whereas effects in vegan dieters were reversed. This underlines the potential confounding of (presumed) expectations towards plant-based meals when being able to deliberately choose between animal-based and plant-based meals. Possibly, food decision-making in non-vegan dieters might not rely on self-set dietary goals as much and reservations about plant-based meals could be further fuelled by an unknown food composition or ingredients or unfamiliar taste. For vegan dieters meal choice is anyhow limited and food decision-making might rely on more long-term goals including health, ethical, and climate arguments ^56^, all of which are not immediately to be processed at time of meal choice. Indeed, literature shows that vegan and vegetarian diets are associated with orthorexia nervosa, an eating disorder with an obsession for healthy eating ^57^. Unexpectedly, in sub-study two with omnivorous dieters only we also found higher scores for cognitive restraint and hunger in those choosing plant-based meals. Overall well-being or emotional constitution were not assessed in the app study though. In sum, dietary adherence factors could have masked or distorted potential effects of meal category on hunger and mood, indicating that other factors than single meal choice exert again considerably stronger influence.

Online studies are gaining momentum, in particular hosted on crowdsourcing platforms, and have great potential in terms of collecting big datasets from the real-world ^58^. Yet, several challenges in our study design should be noted as limitations:

Firstly, all results were based on self-reported subjective ratings and we expected compliance by the participants with the study set-up, yet fake ratings could not be excluded. The virtual setting and a resulting lower commitment to complete the study might also explain relatively large amounts of missing values for some of the confounding variables. Moreover, nutritional values were only available in a subset of data, further limiting power in the respective analyses. Nutritional information is publicly provided by very few providers only (8 out of 57), and was not available upon request. Note, that information on fiber content was not available, which might be due to a general lack of awareness and literacy on dietary fiber (“knowledge fiber gap”, compare ^59^), to which end we advocate to extend nutrient labelling on food packaging, in menus, in nutrition apps and nutritional experimental databases etc. by dietary fiber. Further, online datasets require careful data curation strategies, including exclusion of duplicates, defining outliers, fake and non-compliant entries. Notably, when restricting our analysis for the app-based study to a pre-defined still liberal or conservative timeframe around meal intake when entries needed to have been made, sample sizes were drastically reduced and smaller differences did not reach significance anymore, somewhat limiting our conclusions. Also, while meeting targeted sample sizes in sub-studies 2+3 initially, data curation and exclusion of timepoints and participants led to a higher than expected dropout rates, may have led to underpowered results. To overcome this issue, future app studies might implement a technical solution that reminds participants to stick with certain time frames. Also when cooperating with already existing institutions and technological environments, the research team has to adapt to limitations or peculiarities of the dataset such as missing information or unusable data structures and should thus a priori consider how this would affect statistical analyses. Secondly, Likert scales were conceptualized and adapted to the virtual setting, making the use of established research tools difficult, but instead calling for new solutions and ideas for developing suitable, reliable research tools (such as emoji-based Likert scales). Emojis may be suitable for assessing emotions in humans: perception and interaction with facial emoji Likert scales ^60^ and consumers’ emotional reaction elicited by food or drink products after tasting the products ^61,62^ been investigated previously. Despite assessing a wide range of emojis, extreme anchors were “happy, content, joyful” and “unhappy, dissatisfied, stressed” ^62^. Another, study in children found higher ratings for emoji-based vs. verbal-based Likert scale, yet found emoji-based results to be less reliable and valid ^63^. Overall, the studies show the potential and certain benefits of using emoji-based scales, yet the selection of emojis and the multiple meanings call for considerate application in line with the study design. Further, due to the smartphone-based setting we decided to use subjective momentary well-being as a proxy for mood, refraining from using more comprehensive measures of mood. Thirdly, meal categorisation into animal-based vs. plant-based was based on provided information by the cafeteria meal plans on allergens and meal description using keywords for classification. Manual categorisation was only done in sub-studies 2 and 3. Therefore, only unambiguously classified meals could be included, leading to a reduction in sample size. Furthermore, we did not differentiate between vegetarian and vegan dishes. Fourthly, the timeframe of investigating single meal choices may be a methodological limitation. The present acute effects are somewhat in contrast to epidemiological evidence showing higher satiety and both either higher or lower depressive symptom scores in vegan/vegetarian dieters ^21,23,64^, we did not observe higher satiety and mood at post-meal for plant-based meals. In dietary adherence subgroup comparisons we found that lower satiety and lower mood at post-meal for plant-based choices occurred in the predominantly omnivorous and vegetarian group only, yet effects were inverse for the predominantly vegan subgroup. This could be due to the limited timeframe of the intervention only looking at acute postprandial changes in satiety and mood after a single meal in contrast to long-term dietary habits in epidemiological studies, that might be better suited to capture changes in gut-brain signalling and host physiology in response to higher fiber intake. A systematic review further highlights that although fiber indeed leads to satiety-inducing effects, the expected timeframe of action is rather 3 to 15 hours post-ingestion ^16^. Indeed, it has been shown that diet can rapidly change host gut microbiome, yet these changes only occur within 24 to 72 hours after drastic dietary changes ^65^. Also, we found that individuals choosing a plant-based meal showed lower overall hunger and higher mood before the meal compared to animal-based meals. In sum, these findings underline that contrary to our hypothesis, not the acute meal choice, but rather habitual dietary patterns and other characteristics such as dietary adherence may determine the effects of a meal. Dietary adherence groups are based on self-report and predominant dietary habits, and therefore might not reflect real dietary intake. Note, that self-reported dietary identification may be less distinct for the predominant vegetarian group as compared to predominant omnivores and vegans, due to numerous factors influencing dietary adherence identification in vegetarian dieters ^66,67^. It is estimated that more than 50% of vegetarians were open to eating meat ^66^, therefore our predominantly vegetarian group likely includes semi-vegetarians and flexitarians. Also generalisability to the general population is limited, as our selected sample is a young, student population. Fifthly, we did not assess important psychological factors that might have influenced meal choice, in particular, health awareness, dieting goals, sport habits and ethical concerns.

Besides the listed limitations, the presented studies have several strengths:

Firstly, the real-life experimental setting of all three studies is a major strength aiming to enhance application and conclusions to the living population without being in a highly controlled research environment. Secondly, pre-registered sample size estimations on statistical power were met: especially the app study consists of a huge sample and has been collected nation-wide in Germany and partly in Austria increasing generalizability of the results independent of cafeteria location or regional differences. Also sub-studies 2 and 3, although lower in sample size, are thoroughly phenotyped datasets with a multitude of potential confounders assessed. Indeed, we observed high compliance of the participants and high commitment to submit voluntary pre- and post-meal photos (∼62%). Thirdly, all studies contain pre- and post-measures that allow linear mixed effects modeling correcting for the individual participant and the respective cafeteria location. Fourthly, addressing the complexity of real-life settings, various confounding factors were assessed and included in exploratory analyses to enhance the understanding of multimodal influences on meal decision-making and subsequent hunger, mood and stress ratings. Lastly, all three datasets have been carefully curated and sanity checks have been performed to ensure reliability of virtually collected ratings.

## Conclusion & Outlook

In summary, this series of large-scale surveys showed that overall effects of single plant-based meals compared to animal-based meals on satiety and mood are rather small compared to main effects of timepoint, underlining that meal category only has a minor impact. This notion alleviates some of the reservations against plant-based meals potentially not being satisfying enough or not leading to fullness, in modern societies ^68^. Protein content and taste of meals were shown to contribute to satiety and mood. Thus, they should be increased in university cafeterias, in particular for plant-based meals, in order to enhance satisfaction and lastly also acceptability and prevalence of those meals that are regarded climate-friendly. Increasing plant-based choices remains an important societal task with the aim to improve not only individual but also planetary health ^69^. Moreover, promoting plant-based offers and increasing the proportion of plant-derived ingredients through science communication and the above discussed mechanisms might help to counter the rising obesity epidemic, particularly by replacing Western(-ized) diets.

Overall, animal-based vs. plant-based meal category has only minor influence on post-meal satiety and mood in university cafeterias compared to other demographic and external factors influencing meal choice. Future longitudinal studies, for example using smartphone-integrated real-world big data sampling are warranted to further understand the acute and long-term determinants of healthy food choices, possibly also assessing internal beliefs related to food decision-making. Our findings might help to develop strategies to increase acceptability of healthy and sustainable plant-based food choices.

## Materials and methods

### Ethics

The Ethics Committee of the Medical Faculty of the University of Leipzig approved the study protocol and all participants provided written informed consent. The study was divided into three sub-studies and pre-registered at https://doi.org/10.17605/OSF.IO/A7YTS (28 Nov 2019). All data was collected from Jan 2019 until Feb 2020.

### Recruitment

#### Study 1 - app study

For the first sub-study, all users of the smartphone app iMensa and Mensaplan (provided in Germany, Austria, Switzerland by Aimpulse Intelligent Systems GmbH, Bremen, Germany) were invited to take part in a brief survey consisting of 12 questions. The app has been originally developed to provide meal plan information and collect meal ratings of student cafeterias (mostly run by the local “Studierendenwerk”) with approximately 300,000 users and about 5,000 ratings per week.

#### Study 2+3

*browser-based studies*. In the second and third sub-studies, all visitors of the student cafeterias in Leipzig, Berlin, Halle and Jena who planned to have lunch on-site, were over 18 years old and who had access to a LimeSurvey www.survey3.gwdg.de via a mobile device could take part in the study. Before eating a meal, a baseline survey served as a screening tool to exclude visitors reporting known food allergies, food intolerances, eating disorders, neurological and/or psychiatric disorders, and other diseases that may have an effect on diet and mood like depression, schizophrenia, multiple sclerosis or Crohn’s disease. Also, vegetarians and vegans were excluded for better comparability of meal categories among omnivorous subjects and to enable random allocation to meals for the third sub-study. Participants were free to choose the day of the study.

### Study design

#### Study 1 - app study

In the app, users were asked to report hunger and happiness before and after consuming a meal on a 5-point Likert emoji scale each, accompanied by the originally app-provided rating of the respective meal on a 5-star scale (hunger: *“How hungry are you today before/after meal intake?”* translated from the original *“Wie hungrig bist du heute vor/nach dem Essen?”*; mood: *“How happy are you today before/after meal intake?”* translated from the original *“Wie glücklich bist du heute vor/nach dem Essen?”*). For the additional study-specific questions, emoji-based 5-point scales (**Figure 1**) were used to increase user-friendliness and to blend into the technological interface, selected based on maximal ratings on comprehension of the respective measure of interest in a pilot study (hunger: n = 29, 45% approval for emoji “plate”; mood: n = 30, 47% approval for emoji “smileys”, further details in **SI**). In addition, we asked for meal decision (spontaneous or planned), gender (female, male, diverse), usual predominant dietary habits (omnivorous, vegetarian, vegan) (“*Which dietary pattern do you adhere to predominantly?*”), amount of liquid intake in the last 2 hours (< 0.5L, < 1L, > 1L), whether they ate alone or in company, whether they finished their meal or not, frequent smartphone use during the meal (yes or no), and sleep quality of the last night (good, normal, bad). All data was provided with anonymous user identifiers by the app provider, along with exact date and time stamps of each question. The specifics to the app study (compared to sub-studies 2+3) were 1) both predominantly omnivorous and vegetarians/vegans took part in the study, no non-voluntary meal choice has been performed, 2) mood was asked with “how happy are you today …”?, 3) the study did not assess stress levels and 4) meal categories were specified based on the available information provided by the cafeterias.

#### Study 2+3 – browser-based studies

In sub-studies two and three, after the initial baseline survey, in a subsequent pre-meal survey, participants were asked to rate momentary satiety, happiness and contentment on a 10-point Likert scale scale (hunger: *“How hungry are you right now?”* translated from the original *“Wie hungrig sind Sie im Moment?”*; mood: *“How happy are you right now?”* translated from the original *“Sind Sie im Moment zufrieden?”; stress: “Do you feel stressed right now?”* translated from the original *“Fühlen Sie sich im Moment gestresst?”*) **(Figure 1**, before either freely choosing a meal (sub-study 2) or being randomly assigned to eat a animal-based or a plant-based (vegetarian or vegan) meal (sub-study 3). After finishing the meal, participants were instructed to again report satiety, happiness and contentment on the same 10-point Likert scale. Moreover, age, gender, height (in 5 cm categories) and weight, as well as net monthly income (<500€/500-1000€/1000-1500€/1500-2000€/2000-2500€/>2500€) were assessed. In addition, a) estimated portion size, b) whether the meal was fully eaten up or not (yes/no), (c) whether side dishes or desserts have been chosen, (d) a questionnaire on habitual food intake (Food Frequency Questionnaire (FFQ), DEGS-1 ^70^), e) a personality inventory (NEO-FFI) ^71^ and f) a questionnaire on eating habits (TFEQ) ^48^. In addition, the survey asked about social contact during lunch (Did you have lunch alone or with others? Have you been looking at your mobile longer? Did you sit alone?). Information about the study and its goals was provided right at the beginning of the baseline survey before meal intake. Optionally, participants could upload pre-lunch and post-lunch photos of their meals to monitor meal choice and compliance. In the randomised condition, the post-survey also asked whether participants were satisfied with the randomised condition, or if not, which meal choice they would have preferred.

### Dietary adherence

#### Study 1 - app study

Usual predominant dietary habits (omnivorous, vegetarian, vegan) were asked with *“Which dietary pattern do you adhere to predominantly?”*. Individuals reporting all adherences were eligible to take part in the study.

#### Study 2+3 – browser-based studies

Dietary adherence was asked with *“What is your diet type?”(* translated from the original *“Wie ernähren Sie sich?”*) with the answer options: omnivorous (both plant-based and animal-based), lacto-ovo-vegetarian (incl. dairy products), pesco-ovo-vegetarian (incl. fish and dairy products), pesco-vegetarian (incl. fish), vegetarian, vegan. Only participants choosing omnivorous were admitted to proceed with the study. Based on self-reported Food Frequency Questionnaire data, we estimated mean daily intake of 68 g or weekly intake of 478 g of meat for our selected cohort. German Nutrition Foundation recommendations for meat intake is 300-600g per week. Therefore, we esteem our selected cohort to be within the expected range representative of a standard, German Western-style diet, consuming meat on a regular basis.

### Remuneration

#### Study 1 - app study

Participants of sub-study 1 were not remunerated due to a very brief study completion time.

#### Study 2+3 – browser-based studies

Sub-studies 2+3 were remunerated with 9 € for participation, with an optional extra 2 € per questionnaire (TFEQ, FFQ, NEO-FFI).

### Measures

#### Main variables

As pre-registered, main outcome measures were mood (pre/post), satiety (pre/post), stress (pre/post) (only in studies 2+3). As preregistered, we omitted stress levels in the app-based study due to anticipated short attention span whilst using smartphone-embedded surveys and to maximize re-voting on multiple days on our two main questions which are mood and hunger. For this project, mood is defined as subjective momentary well-being, capturing happiness ^72^, and can therefore only be considered as a proxy for mood, as no comprehensive scales or validated questionnaires have been employed.

#### Control variables: Study 1 - app study

Control variables included meal category, time lag between measures, macronutrients (calories kcal / 100g, carbohydrates mg / 100g, sugar mg / 100g, fat mg / 100g, saturated fats mg/100g, protein in mg / 100g), fluid intake in last 2h (<0.5l/<1.0l/>1.0l), predominant dietary adherence (omnivorous/vegetarian/vegan), gender (f/m/d), sleep (bad/normal/good), social interaction during meal (alone/in company), prolonged mobile use during meal (yes/no), finished meal (yes/no), pre-meal planning (planned decision/impulsive decision), liking of meal (taste rating from 1-5 stars), place of meal (name of cafeteria).

#### Control variables: Study 2+3 – browser-based studies

Control variables included meal category, time lag between survey responses, age (), gender (f/m/d), body size and body weight to compute BMI in kg/m^2^, net monthly income (<500€/500-1000€/1000-1500€/1500-2000€/2000-2500€/na), estimated portion size (very small/small/normal/big/very big), fully eaten up or not (yes/no), drinking volume last 2h (water, sugar drinks, tea, coffee, juice, alcoholic beverages, others; each on a 6-point scale), side dishes (yes/no and meal description) or desserts (yes/no and meal description), social interaction during meal (alone/ in company), prolonged mobile phone use during meal (yes/no). Questionnaires on well-being (WHO-5 ^73^), eating traits (TFEQ ^74^), personality traits (NEOFFI ^71^) and regular eating habits (DEGS-1 FFQ ^70^) were administered. Questionnaires were scored according to the respective manual. For FFQ data additional scoring on food intake quantity was performed to retrieve nutrient intake as described previously ^75^.

### Meal categorisation

For analysis meal categorisation was done by study personnel based on provided meal category (vegetarian, vegan, meat or fish) and more specific meal description as given in the menu and by information on allergens (keywords: *animal-based*: poultry, pork, beef, lamb, meat, fish, scampi, seafood, deer, piglet, duck, goose, salami, ham, wiener; *plant-based*: vegan, vegetarian, meat-free, tofu, soy). In brief, categorisation was done so that animal-based meals contained animal flesh and plant-based meals did not, the latter consisting of both vegetarian and vegan meals. Categorisation was done manually and in case of ambiguity based on consensus amongst study personnel. All remaining ambiguous meals were assigned missing value (e.g. in case meal description included both animal-based and plant-based keywords, e.g. “Tofuwurst oder Currywurst”, “…also available in vegan”, “Salatbuffet je 100g Auswahl an frischen Salaten, Gemüse-, Fisch-, Geflügel-, Schweine- und Rindfleisch- und vegetarischen und veganen Gerichten”).

### Missing data and data selection/exclusion

#### Plausibility

All data was screened for plausibility, consistency and distribution and curated as appropriate. This included assigning meal category based on available information (meal description, information on allergens, meal photo). In sub-study 3, non-compliance with the assigned meal category led to exclusion. Implausible datapoints were treated as missing. Missing data was treated as missings (linear mixed models), except for meal category, which led to exclusion of the datapoint in all analyses.

#### Time stamps

Data entry plausibility was monitored with time stamp information (app study) and, if available, based on the creation dates of the uploaded meal photos (substudies 2+3). In sensitivity analyses, data was excluded according to the following criteria: post-meal entries appeared to be made prior than pre-meal entries; data entry outside of liberal time frame for more than 5 min and more less 3 h time lag between pre- and post-meal (hunger) entries (pre- and post-meal entries outside of 7:45 am and 10:30 pm (cafeteria opening hours (10:45 am to 7:30 pm) + 3 h data entry time frame); as well as data entry outside of a conservative time frame more than 20 min and less than 1.5 h time lag between pre- and post-meal (hunger) entries; (pre- and post-meal entries outside of 9:15 am and 9 pm (cafeteria opening hours + 1.5 h data entry time frame)) (note: we corrected timings indicated in the pre-registration). In sub-studies 2 and 3 only, duplicate records of the same participant were checked for consistency and curated accordingly so that each participant would only be counted once.

For detailed information on data curation for sub-studies 2+3, see https://osf.io/ucq6r/.

### Data analysis

As pre-registered (https://doi.org/10.17605/OSF.IO/A7YTS), main pre-registered *s*tatistical analyses comprised linear mixed model comparisons using lmer() in R for hunger, mood and stress as dependent variables, meal and timepoint as factors and/or interaction terms, modelling individual and cafeteria as random factors. Alpha level was adjusted to multiple comparisons (3 main tests per study), with the significance threshold set to p < 0.05/3 = 0.0167, and p < 0.05 for all other (and exploratory) models. For pre-registered secondary analyses, chi-square tests were performed for categorical outcome (planned vs. impulsive decision). Deviating from the pre-registration, models (and planned mediation analyses) for fiber content *<u>could not be performed</u>*, as data was not available. All other statistical analyses, including subgroup analysis and correlation analysis, are considered exploratory analysis.

### Power calculation

The ethics proposal submitted to the institutional ethics board of the Medical Faculty of the University of Leipzig, Germany, (228/18-ek) included power calculations for all three studies.

#### Study 1 - app study

For the app study, power calculation was based on a study by Koenig et al. (2018), resulting in a target sample size of 126, assuming the observed effect size. This study observed an effect in the eating decision behavior depending on the adaptation of a behavior model (η^2^ = 0.03, corresponds to an effect size d = 0.35 (https://www.psychometrica.de/effect_size.html)). However, it is difficult to estimate the effect size of our study, since many influencing factors cannot be determined within the framework of an app study. The analyses therefore remain largely explorative. Due to the large number of users of the app (currently: approx. 300,000 users, approx. 30,000 meal evaluations per month), groups of the same size can be pseudorandomized for better statistical evaluation. If 10% of active users are included in the study, 3,000 meal-well-being associations per month can be collected. Therefore, we assume to achieve a sufficient sample size to determine a possible effect.

#### Study 2+3 – browser-based studies

Sub-studies 2 + 3 were based on the effect size of the study by Reed and Ones (2006) (d = 0.47), which examined the effect of a one-time sporting activity on mood values (studies that examined the influence of a one-time meal on mood are missing so far). Study 2 has the following special features: a) a smaller effect was assumed (d = 0.30) than in study 3, because of the absence of randomization, b) a dropout rate of 20% was added to the calculated sample size, c) a 1:3 distribution from plant-based dishes to meat dishes was assumed. Thus, based on the sample size of the power analysis of 172 test people, a sample size of 210 subjects suggested to observe the desired effect. For sub-study 3, power calculation is analogous to sub-study 2, but defers in the following points: a) effect size of the study by Reed and Ones (2006) was assumed (d = 0.47), which investigated the effect of a one-time physical activity intervention on mood. Assuming a dropout rate of 20%, we assume that a targeted sample size from 85 (up to 6,300 portions per day in one of the locations of the Leipzig student union, source: Mensa im Park, see https://www.l-iz.de/detector/movement-detector/2015/04/three-headed-team-takes-over-leading-of-the-mensa-am-park-des-studentenwerkes-leipzig-84262).

### Stopping rule

#### Study 1 - app study

Sample size in the app-study largely exceeded the predicted sample of n = 126 to detect a small effect (n = 16,135 unique ratings). This was indeed due to the rapid data collection within the app and data extraction after certain time periods as agreed on in the contract with the developer. Therefore, during data collection, we could not monitor exact ratings, but rather stopped data collection after the given time frame.

#### Study 2+3 – browser-based studies

For sub-studies 2+3, estimated sample sizes were strictly adhered to while recruiting in person and stopped once the expected numbers plus estimated drop-out rates were obtained. We met the targeted sample size, yet data curation and exclusion of timepoints and participants led to a higher than expected dropout rate and therefore, sample size was not met in sub-study 3.

### Meal photos

#### Study 2+3 – browser-based studies

Meal photos before and after meal intake were optionally uploaded by the participants in sub-studies 2+3 (before data curation 78% (sub-study 2: 216 out of 273; sub-study 3: 114 out of 149)), after data curation 62% (sub-study 2: 108 out of 173; sub-study 3: 44 out of 71) with pre- and post-meal photos). Photos were used to check accordance with self-reported meal description, as well as time stamps of the photos and compliance with study design for randomized choice in sub-study 3. All photos are stored at https://osf.io/mqc5d/ and can be used for further nutrient analysis or other purposes upon request to the corresponding author.

### Emoji-based Likert scales

*Study 1 - app study*. Due to the absence of suitable, validated emoji-based scales, we self-developed those and validated comprehension of the measure of interest in a pilot study in n ≥ 26. (**Supplementary Figure 1**). We showed participants (researchers in the fields of neuroscience, psychology, medicine) scales and let them rate two dimensions. Firstly, *“What do you think is most likely to be rated on this scale?”* with an open text field, and second: *“Which scale (listed from top to bottom 1 to 3) represents “momentary well-being” best?”* (or the respective construct) in a rank order question compared to all other scales representing this construct. We then quantified relative preference ratings for each construct, and chose those scales, that showed highest relative preference for representing a construct.

## Data Availability

Raw data, scripts and meal photos are stored at https://osf.io/mqc5d/ and can be used for further nutrient analysis or other purposes upon request to the corresponding author.

## Data availability

The datasets generated and analysed during the current study are available in the Open Science Framework repository at https://osf.io/mqc5d/.

## Code availability

The code generated during the current study are available from the corresponding author on reasonable request.

## Acknowledgments

We would like to thank Maria Paerisch for the administrative advice and support for setting up the online studies, Lena Weidert for initial data curation of sub-studies two and three and David Villringer for initial data analysis of the app study. We thank Aimpulse Intelligent Systems GmbH, Bremen, Germany and the local Studierendenwerke (student unions) for integrating our study into their services and for providing the data. We also thank all the participating volunteers. Funding was provided by the Max Planck Society and a promotion stipend by the German Federal Environmental Foundation (EM). Open Access funding enabled and organized by Projekt DEAL.

## Author Contributions

EM, AV, AVW: study conception and design; EM, LdB, MZ: data collection and data management; EM: data curation; AVW: meal category labelling for the app-study; EM, AVW: data analysis of all sub-studies; EM: data visualization; EM, AVW: first manuscript draft; EM: revisions. All authors contributed to and accepted the final manuscript.

## Figure Legends

Table 1: Demographic information across studies according to meal choice. NB: Significant differences according to Chi-tests or Wilcoxon tests between meal categories are marked in bold. Abbreviations: N.A.: no answer; F: female; D: diverse; M: male

Table 2: Ratings for hunger, mood, stress across all studies. Mean ± SD. Significant models of interest compared to null model in bold (LMM comparison p<0.001). NB: Significant effects between linear mixed models model comparisons are marked in bold. Note, that due to multiple comparison correction alpha-level was set to 0.05/3 = 0.0167. P-values represent ANOVA model comparison of linear mixed models with fixed and random effects.

