## Supplementary Information for "Effects of single plant-based vs. animal-based meals on satiety and mood in real-world smartphone-embedded studies"

### Sample descriptives

*Study 1 - app study.* In the app-based study, planned meal decisions were more frequently made for plant-based (72%) compared to animal-based meals (62%). Eating alone was more frequent (23%), when having a plant-based compared to animal-based meal (21%). Finishing the chosen meal was slightly more frequently reported by individuals choosing an animal-based (87%) compared to plant-based meal (86%). Drinking volume before the meal, smartphone use and sleep was not different across meal categories.

*Study 2+3 – browser-based studies.* In sub-studies 2 and 3, neither body mass index (BMI) nor income nor general well-being differed across meal categories. Additionally, in sub-study 2 omnivores choosing a plant-based meal were characterized by similar habitual nutrient intake compared to omnivores choosing animal-based meals, except for significantly higher fiber intake (22 ± 11 vs. 18 ± 9 g fiber / day) and significantly higher scores for cognitive restraint and hunger in the TFEQ subscales, **Table 1**). No differences in personality traits or general well-being were found. Respective differences were not significant for the randomized allocation samples in sub-study 3.

### Meal choice descriptives

Meals chosen by category were between 47-61% animal-based; with 55% animal-based in the app-study (8848 animal-based, 7287 plant-based) 61% animal-based in sub-study two (105 animal-based, 68 plant-based) and 47% animal-based in sub-study three (37 animal-based, 41 plant-based).

### Subgroup analysis - Dietary habits (study 1)

To account for potential effects of dietary adherence, we repeated the analysis in subgroups for predominantly omnivorous (n = 11,600), predominantly vegetarian (n = 3,456) and predominantly vegan (n = 911) dieters.

The analysis in predominantly omnivorous dieters only presented the same effects as the whole group, i.e. showing slightly lower mood after a plant-based meal compared to animal-based meals, and also significantly higher post-meal hunger ratings for those choosing plant-based meals (**Supplementary Table 4**). Influences of taste remained significant for post-meal mood, yet the influence of taste ratings on hunger by meal category was no longer significant.

For predominantly vegetarian dieters only, results again resembled the main analysis, with no effect on post-meal hunger, yet lowered mood for those choosing a plant-based meal (note the unequal group size of animal-based meal n = 577, plant-based meal n = 2,880, **Supplementary Table 4**). When accounting for taste ratings, post-meal hunger was lower overall for meals rated with five stars, yet hunger was higher when this was a plant-based meal. Mood ratings showed a similar paradoxical effect when adjusted for taste, namely improved mood for liked meals, yet when those were plant-based, mood was lowered.

For predominantly vegan dieters only, hunger was lower and mood was higher after choosing a plant-based meal (note the unequal group size of animal-based meal n = 117, plant-based meal n = 795 out of 911 vegan dieters, **Supplementary Table 4**). When accounting for taste, highly liked meals reduced hunger, yet there was no difference in hunger ratings between meal categories. Mood adjusted for taste, showed improved mood for highly liked meals, which was also shown for plant-based meals compared to animal-based ones.

Taste ratings by meal category differed for dietary adherence groups, namely that animal-based meals were rated higher compared to plant-based meals by omnivores (animal-based: 3.87±1.2, plant-based: 3.70±1.3) and similarly, but less pronounced for predominantly vegetarians (animal-based: 3.88±1.2, plant-based: 3.81±1.7). The opposite was true for predominantly vegans (animal-based: 3.34±1.7, plant-based: 3.96±1.2) (**Supplementary Figure 9**).

### Subgroup analysis - Time stamps (study 1)

To restrict entries to a reasonable timeframe around actual food intake, we additionally restricted the analyses as preregistered to entries that had more than 5 min and up to 3 h time lag between pre- and post-meal (hunger) entries (liberal timeframe) or more than 20 min and up to 1.5 h time lag between pre- and post-meal (hunger) entries (conservative timeframe). This led to a profound drop in sample size (liberal n = 3725, conservative n = 1878).

Results remained largely unchanged for the liberal timeframe from 5 min to 3 h difference between pre- and post-meal entries, namely significant main effects of post-meal timepoint with lower hunger and higher mood. For interaction effects of post-meal by meal category, no significant effects were found for hunger or mood, yet nominal differences were similar to the main analysis for hunger (higher post-meal hunger for plant-based meals), but not for mood (higher post-meal mood for plant-based meals). For the more conservative timeframe from 20 min up to 1.5 h post-meal, main effects for lower hunger and higher mood for post-meal timepoint were sustained. Yet, interaction effects of timepoint by meal category showed significant higher post-meal hunger for those choosing a plant-based meal ($b = 0.14$, $t = 2.3$, $p = 0.024$), while the main analysis showed non-significant results in the same direction of the estimate, and no significant effect on post-meal mood, yet also similar nominal differences with lower mood after plant-based meals ($p > 0.14$). Note that the reduced sample size when curating for timeframe indicate that the remaining entries had quite a high variety of time lag related to meal intake. Compliance with study design specific timeframes was manually curated in sub-studies 2 and 3 and have already been considered in all of the above analyses.

### Confounder analysis – Coffee intake (studies 2+3)

Although not different across meal categories, coffee intake was assessed as a potential confounder on satiety and mood ratings in sub-studies two and three (data was in subgroups only and not available for the app study). Coffee intake did not explain a significant variance on hunger or contentment, but higher coffee consumption was related to higher stress levels (250-500ml: $b = -0.17$, $t = -0.3$; 500-1000ml: $b = 6.2$, $t = 3.6$; note the limited sample size of n = 32)

### Supplementary Figure Legend

Supplementary Figure 1: Overview of pilot study for validating the self-developed emoji-based Likert scales.

Supplementary Figure 2: Flowchart with sample size for all studies for all sub-analyses.

Supplementary Figure 3: Frequency of meal category choice plotted by federal states across Germany (colour coding by local student union) for the app study. More detailed data available upon request. A: Berlin B: Saxony C: North-Rhine Westphalia D: Thuringia E: Bavaria F: Baden Württemberg G: Sachsen Anhalt H: Bremen I: Hamburg J: Hesse K: Saarland L: Brandenburg M: Mecklenburg Western Pomerania N: Lower Saxony O: Rhineland Palatinate P: Schleswig Holstein.

Supplementary Figure 4: Participation numbers plotted by city and by meal category in time (studies 2+3).

Supplementary Figure 5: Eating behaviour of individuals in sub-studies 2+3 for A) nutrient intake based on 1-week FFQ data and B) eating traits according to Three-Factor Eating Questionnaires subscales.

Supplementary Figure 6: Word clouds and frequency tables of words in meal description information for A) animal-based category and B) plant-based category from the app study. Word clouds created with *https://tagcrowd.com/* and frequency tables with *https://countwordsfree.com/*

Supplementary Figure 7: Correlation of post-pre changes between hunger, mood and stress levels for sub-studies 2 and 3. Spearman's correlation and 99.9% CI.

Supplementary Figure 8: Frequency of taste ratings of meals per meal category per gender (app study only).

Supplementary Figure 9: Frequency of taste ratings of meals per meal category per dietary adherence group (app study only).

Supplementary Table 1: Macronutrient composition differences between meal categories ($n_{max}$ = 1262, data from app study).

Supplementary Table 2: Interaction effects of taste ratings on hunger and mood post-meal (app study only).

Supplementary Table 3: Interaction effects of meal category on hunger and mood for subgroups by gender (app study only).

Supplementary Table 4: Interaction effects of meal category on hunger and mood for subgroups according to dietary adherence (app study only).

| Construct of interest | Mood | Hunger | Amount of consumed water | Type of meal decision | Social interaction during meal |
|---|---|---|---|---|---|
| Results (absolute preference votings for each scale and most preferred scale) | 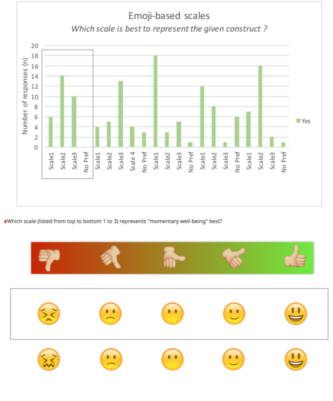 | 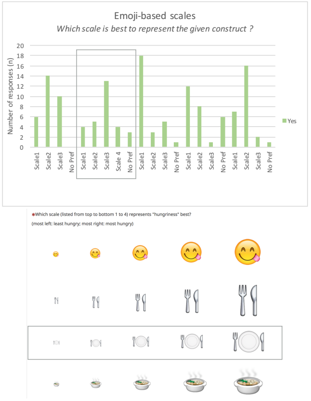 | 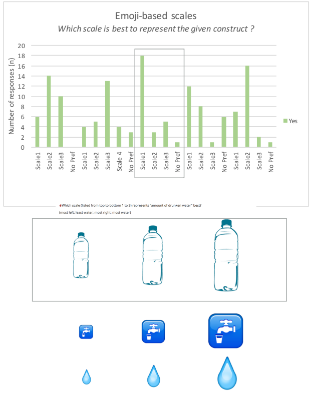 | 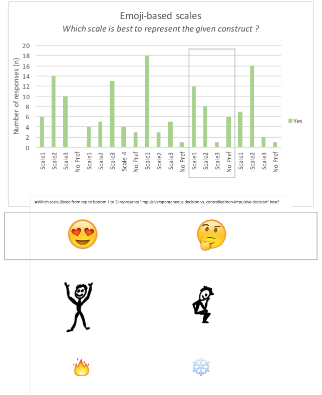 | 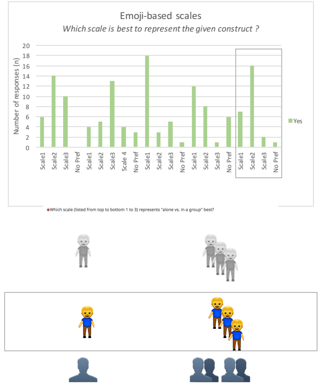 |
| Preference of scale for representing the construct best compared to other scales | Scale 1: 20% (6 out of 30)<br>**Scale 2: 47% (14 out of 30)**<br>Scale 3: 33% (10 out of 30)<br>No Preference: 0% (0 out of 30) | Scale 1: 14% (4 out of 29)<br>Scale 2: 17% (5 out of 29)<br>**Scale 3: 45% (13 out of 29)**<br>Scale 4: 14% (4 out of 29)<br>No Preference: 10% (3 out of 29) | **Scale 1: 67% (18 out of 27)**<br>Scale 2: 11% (3 out of 27)<br>Scale 3: 19% (5 out of 27)<br>No Preference: 4% (1 out of 27) | **Scale 1: 44% (12 out of 27)**<br>Scale 2: 30% (8 out of 27)<br>Scale 3: 4% (1 out of 27)<br>No Preference: 22% (6 out of 27) | Scale 1: 27% (7 out of 26)<br>**Scale 2: 62% (16 out of 26)**<br>Scale 3: 8% (2 out of 26)<br>No Preference: 4% (1 out of 26) |

Supplementary Figure 1: Overview of pilot study for validating the self-developed emoji-based Likert scales.

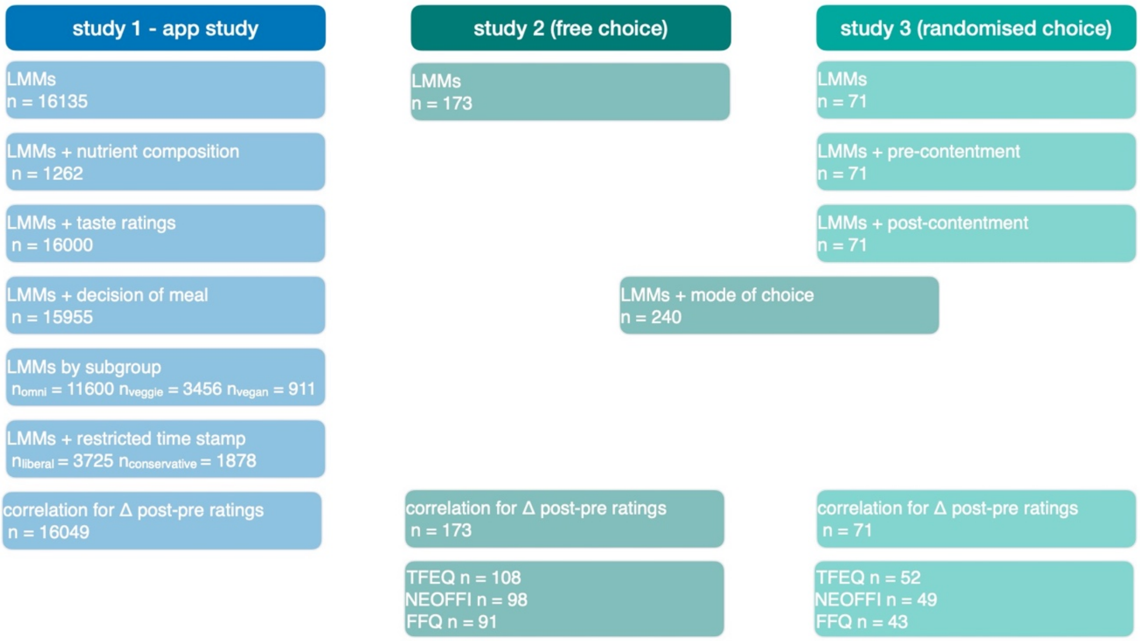


Supplementary Figure 2: Flowchart with sample size for all studies for all sub-analyses.

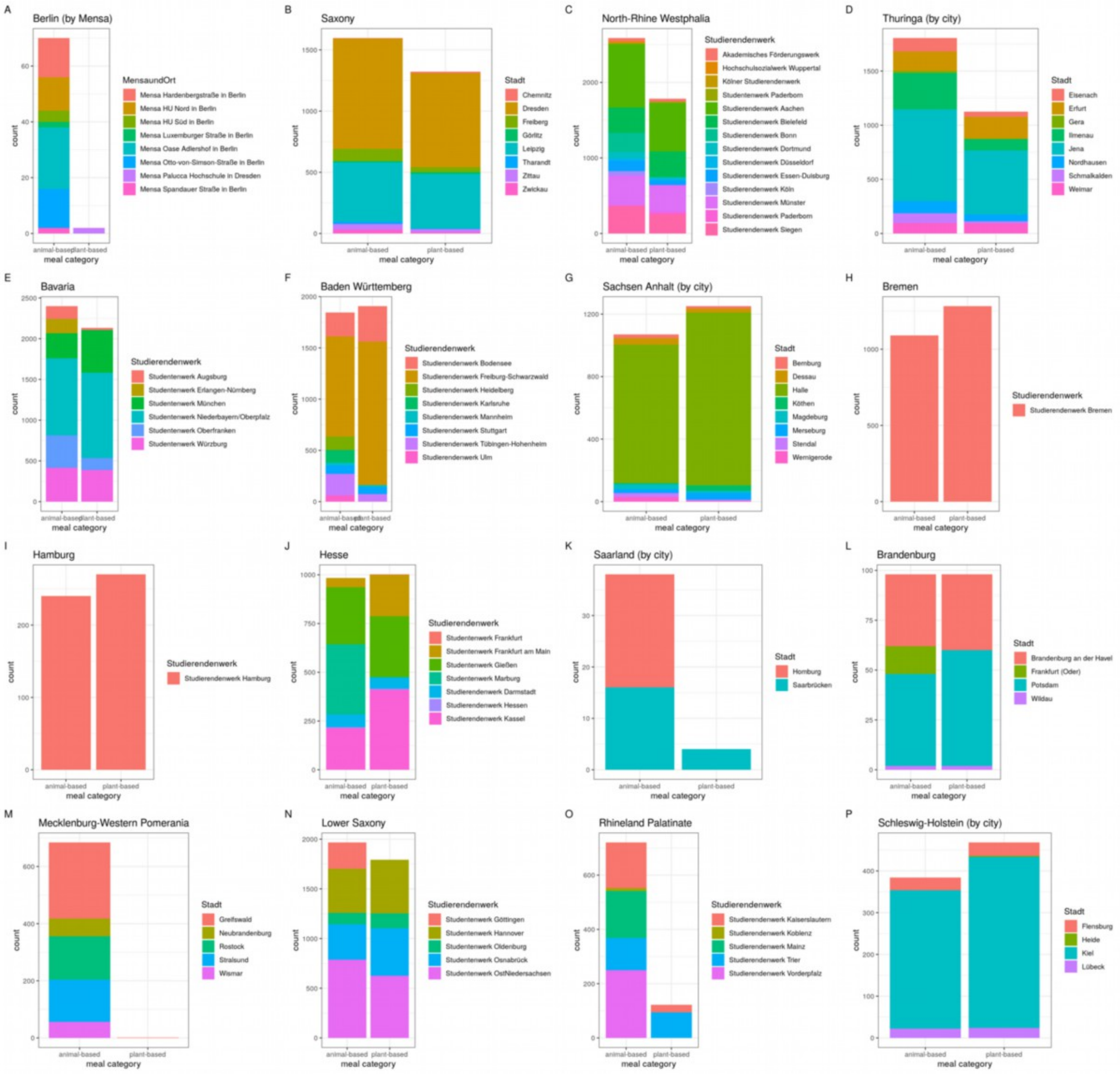


Supplementary Figure 3: Frequency of meal category choice plotted by federal states across Germany (colour coding by local student union) for the app study. More detailed data available upon request. A: Berlin B: Saxony C: North-Rhine Westphalia D: Thuringia E: Bavaria F: Baden Württemberg G: Sachsen Anhalt H: Bremen I: Hamburg J: Hesse K: Saarland L: Brandenburg M: Mecklenburg Western Pomerania N: Lower Saxony O: Rhineland Palatinate P: Schleswig Holstein.

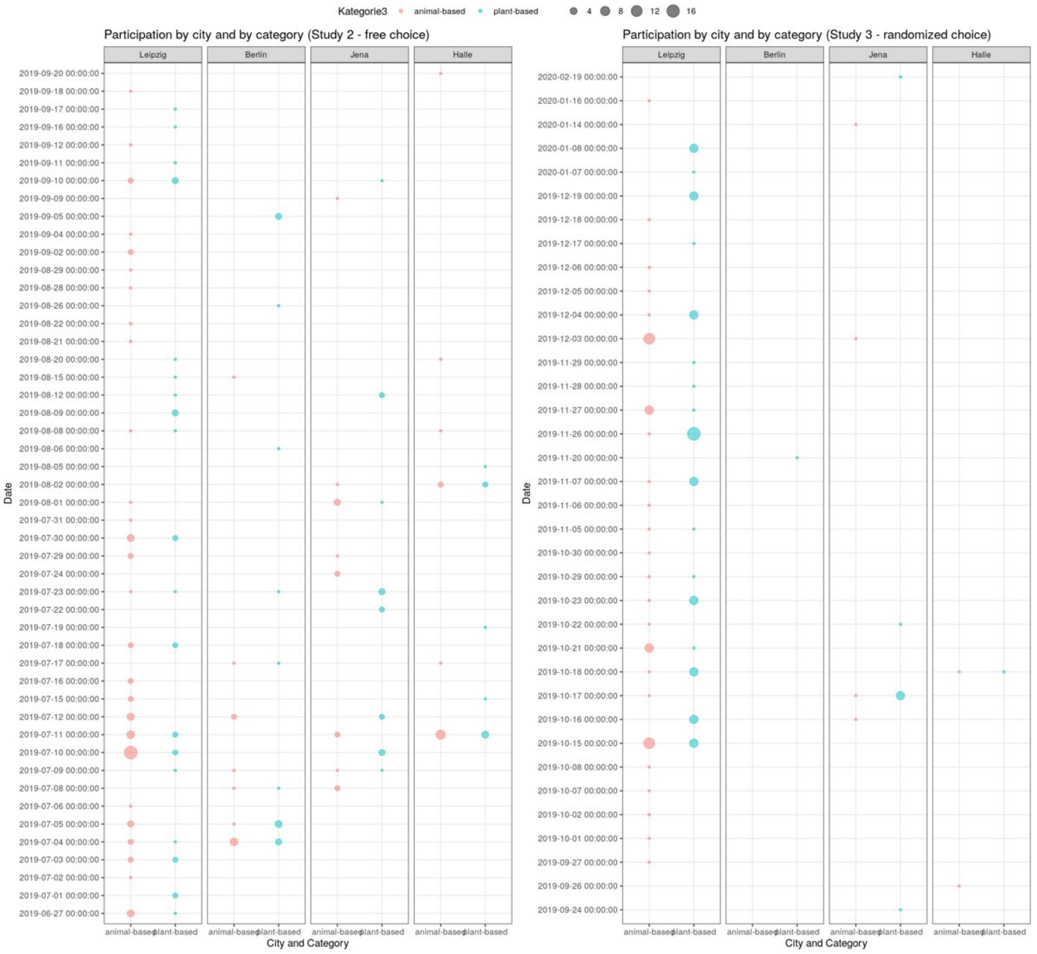


Supplementary Figure 4: Participation numbers plotted by city and by meal category in time (studies 2+3).

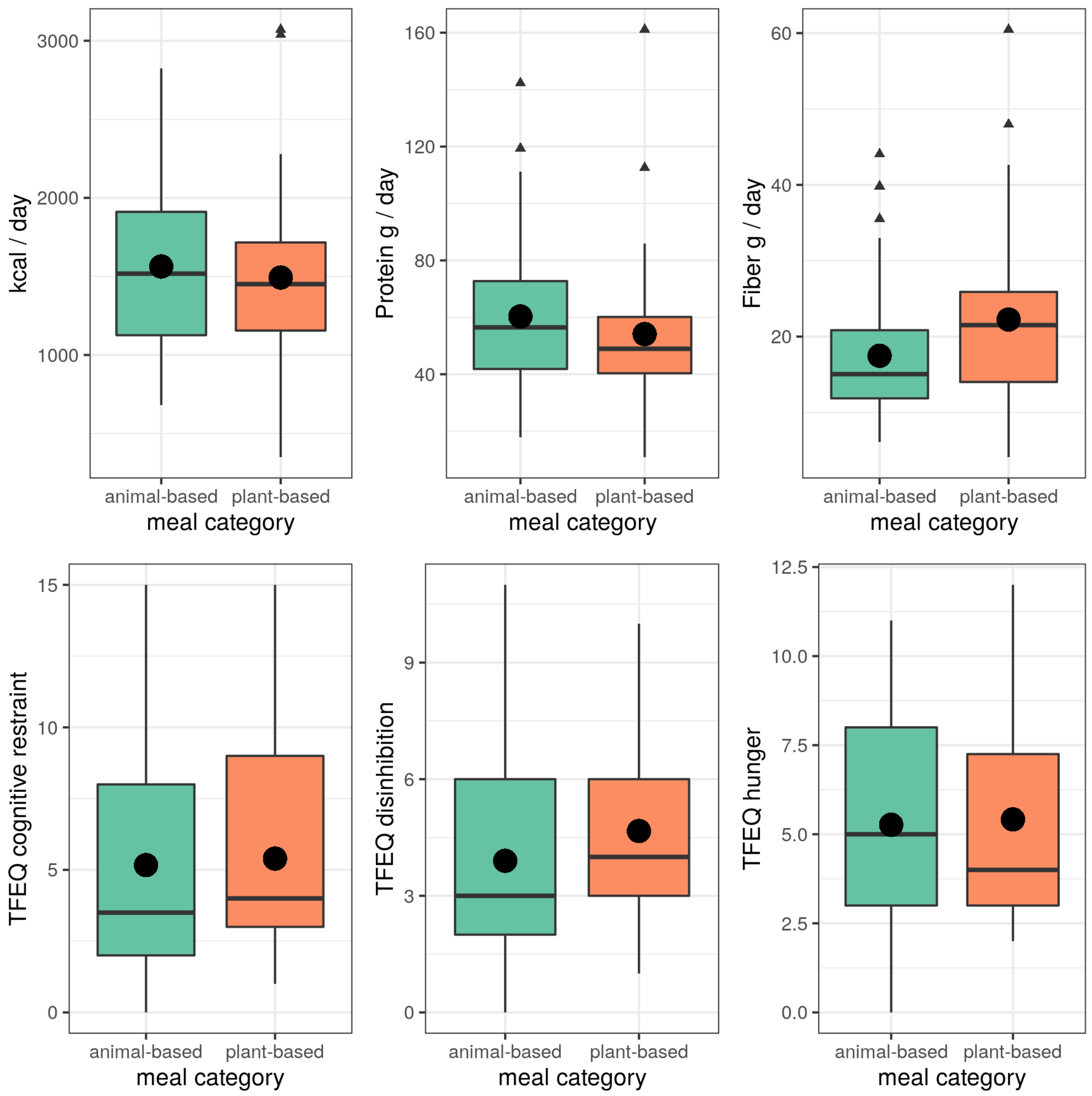


Supplementary Figure 5: Eating behaviour of individuals in sub-studies 2+3 for A) nutrient intake based on 1-week FFQ data and B) eating traits according to Three-Factor Eating Questionnaires subscales.

A)

| Rank | Word | Count | % of text | symbols |
|---|---|---|---|---|
| 1 | Fries (“pommes”) | 709 | 0.8% | 6 |
| 2 | Fries (“frites”) | 700 | 0.8% | 6 |
| 3 | Soup of the day (“Tagessuppe”) | 393 | 0.8% | 10 |
| 4 | Pork schnitzel (“Schweineschnitzel”) | 379 | 1.3% | 17 |
| 5 | Salad (“Salat”) | 343 | 0.3% | 5 |
| 6 | Chicken breast (“Hühnchenbrust”) | 313 | 0.8% | 13 |
| 7 | Rice (“Reis”) | 284 | 0.2% | 4 |
| 8 | Vegetables (“Gemüse”) | 267 | 0.3% | 6 |
| 9 | Breaded (“Paniertes”) | 266 | 0.5% | 9 |
| 10 | Sauce (“Sauce”) | 262 | 0.3% | 5 |

B)

| Rank | Word | Count | % of text | symbols |
|---|---|---|---|---|
| 1 | vegetables (“Gemüse”) | 409 | 0.6% | 6 |
| 2 | pasta (“pasta”) | 258 | 0.3% | 5 |
| 3 | Salad (“Salat”) | 241 | 0.3% | 5 |
| 4 | Tomatoes (“Tomaten”) | 172 | 0.3% | 7 |
| 5 | Side salad (“Beilagensalate”) | 160 | 0.6% | 14 |
| 6 | Rice (“Reis”) | 159 | 0.2% | 4 |
| 7 | Chosen (“gewählt”) | 158 | 0.3% | 7 |
| 8 | Optional side dishes (“Wahlbeilagen”) | 158 | 0.5% | 12 |
| 9 | Bell Pepper (“Paprika”) | 156 | 0.3% | 7 |
| 10 | Mozzarella (“Mozzarella”) | 151 | 0.4% | 10 |

Supplementary Figure 6: Word clouds and frequency tables of words in meal description information for A) animal-based category and B) plant-based category from the app study. Word clouds created with *https://tagcrowd.com/* and frequency tables with https://countwordsfree.com/.

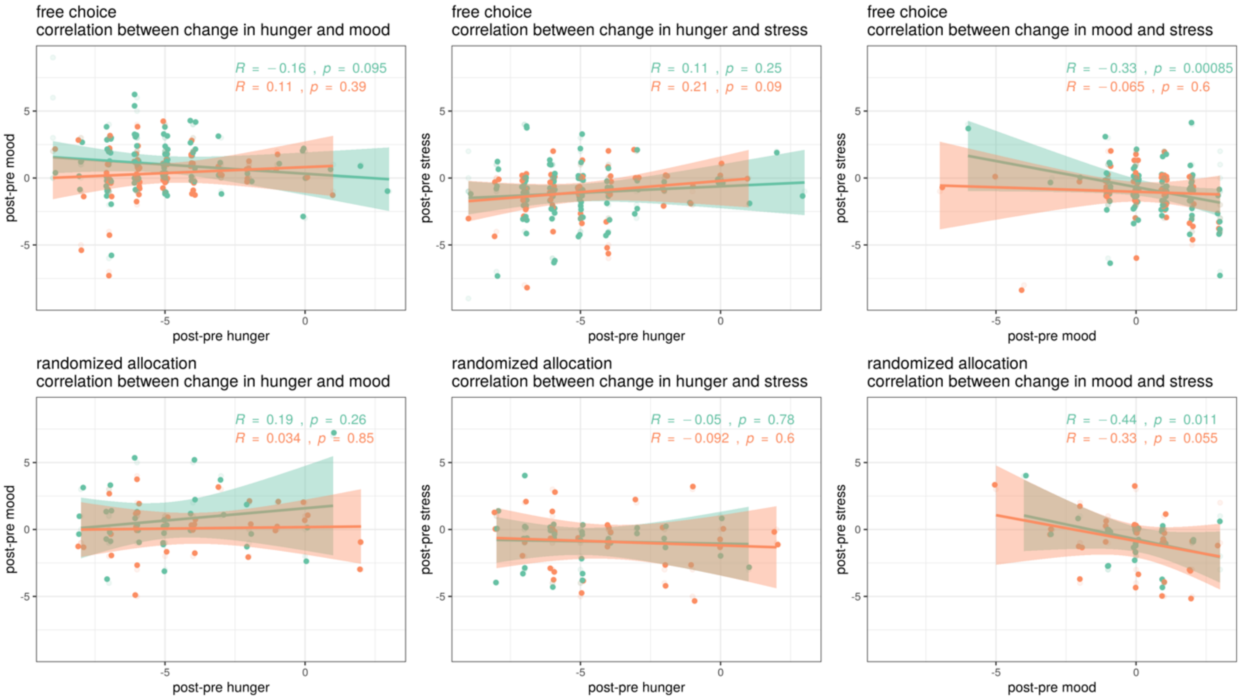


Supplementary Figure 7: Correlation of post-pre changes between hunger, mood and stress levels for sub-studies 2 and 3. Spearman's correlation and 99.9% CI.

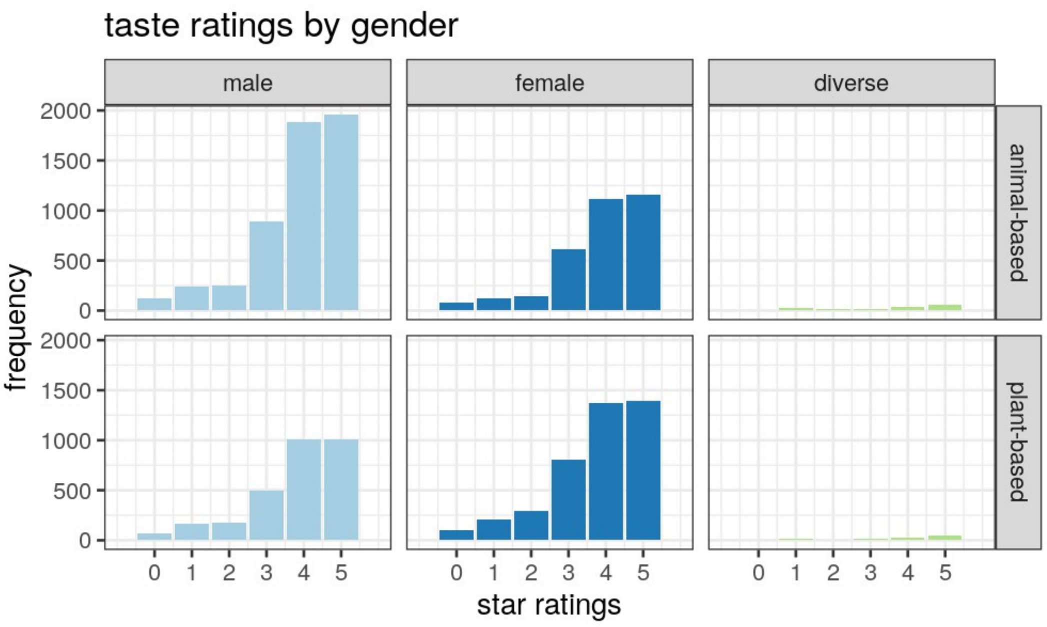


Supplementary Figure 8: Frequency of taste ratings of meals per meal category per gender (app study only).

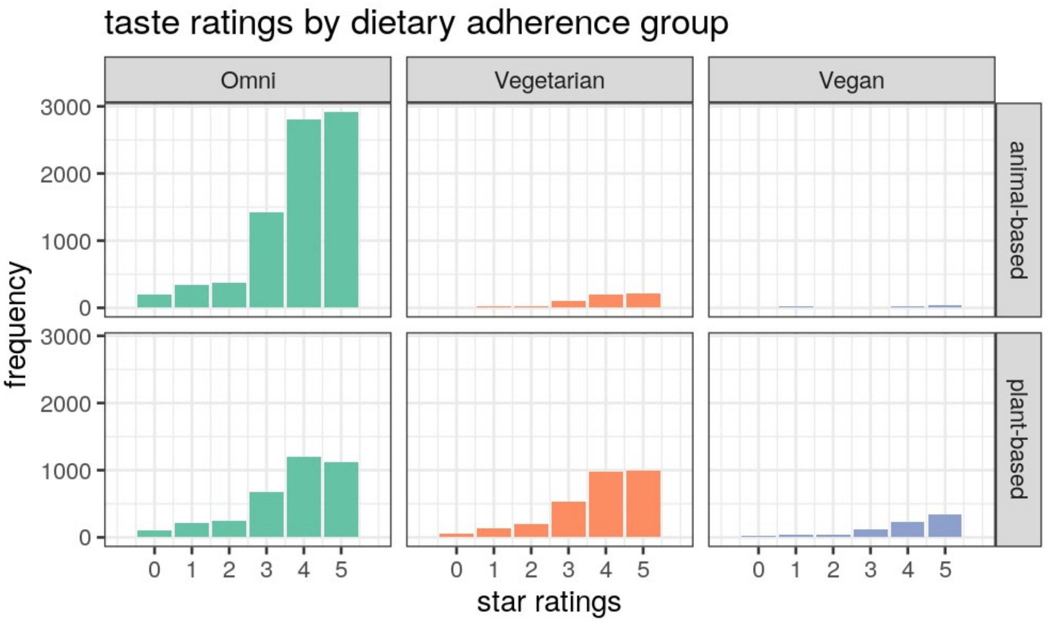


Supplementary Figure 9: Frequency of taste ratings of meals per meal category per dietary adherence group (app study only).

Supplementary Table 1: Macronutrient composition differences between meal categories ($n_{max}$ = 1262, data from app study).

| Nutrient | animal-based meal mean ± SD | plant-based meal mean ± SD | Wilcoxon p-value |
|---|---|---|---|
| **Energy [kcal]** | 591 ± 261 | 572 ± 251 | 0.34 |
| **Carbohydrates [g]** | 50 ± 38 | 71 ± 38 | **<0.001** |
| **Sugar [g]** | 8 ± 5 | 12 ± 11 | **0.002** |
| **Fat [g]** | 28 ± 19 | 23 ± 16 | **<0.001** |
| **Saturated fats [g]** | 8 ± 7 | 8 ± 7 | 0.86 |
| **Protein [g]** | 34 ± 13 | 18 ± 10 | **<0.001** |
| **Fiber [g]** | na | na | |

*Significant differences according to Wilcoxon tests between meal categories are marked in bold.*

Supplementary Table 2: Interaction effects of taste ratings on hunger and mood post-meal (app study only).

| | **hunger** | | **mood** | |
|---|---|---|---|---|
| **app-based (5-point)<br>n = 16000 valid** | **animal-based** | **plant-based** | **animal-based** | **plant-based** |
| **incl. taste ratings** | | | | |
| double interaction effect<br>tp*taste | Fixed effects:<br>Estimate Std. Error t value<br>(Intercept) 3.928288 0.056690 69.294<br>timepointpost -1.686856 0.070519 -23.921<br>meal_cat_corr_2021VEG*N -0.107515 0.017840 -6.027<br>Sterne_rep1 -0.087306 0.067213 -1.299<br>Sterne_rep2 -0.247315 0.066097 -3.742<br>Sterne_rep3 -0.263473 0.058619 -4.495<br>Sterne_rep4 -0.154983 0.056920 -2.723<br>Sterne_rep5 0.032319 0.056861 0.568<br>timepointpost:meal_cat_corr_2021VEG*N 0.005401 0.021959 0.246<br>timepointpost:Sterne_rep1 0.548135 0.085418 6.417<br>timepointpost:Sterne_rep2 0.128536 0.083982 1.531<br>timepointpost:Sterne_rep3 -0.150875 0.074454 -2.026<br>timepointpost:Sterne_rep4 -0.380888 0.072315 -5.267<br>timepointpost:Sterne_rep5 -0.526199 0.072237 -7.284<br><br>**post-meal*5stars: b = -0.53, t = -7.3**<br><br>**$p < 2.2 \times 10^{-16}$** | | Fixed effects:<br>Estimate Std. Error t value<br>(Intercept) 3.360827 0.049121 68.419<br>timepointpost 0.145961 0.052293 2.791<br>meal_cat_corr_2021VEG*N 0.054051 0.015448 3.499<br>Sterne_rep1 -0.002535 0.057777 -0.044<br>Sterne_rep2 -0.065526 0.056820 -1.153<br>Sterne_rep3 0.017549 0.050397 0.348<br>Sterne_rep4 0.133737 0.048933 2.733<br>Sterne_rep5 0.186256 0.048883 3.810<br>timepointpost:meal_cat_corr_2021VEG*N -0.023818 0.016240 -1.467<br>timepointpost:Sterne_rep1 -1.251575 0.063366 -19.752<br>timepointpost:Sterne_rep2 -0.695215 0.062218 -11.174<br>timepointpost:Sterne_rep3 -0.135059 0.055191 -2.447<br>timepointpost:Sterne_rep4 0.179681 0.053613 3.351<br>timepointpost:Sterne_rep5 0.454303 0.053555 8.483<br><br>**post-meal*5stars: b = 0.45, t = 8.5**<br><br>**$p < 2.2 \times 10^{-16}$** | |
| triple interaction effect<br>tp*meal_cat*taste | Fixed effects:<br>Estimate Std. Error t value<br>(Intercept) 3.971202 0.074293 53.453<br>timepointpost -1.827273 0.093326 -19.579<br>meal_cat_corr_2021VEG*N -0.204814 0.110620 -1.852<br>Sterne_rep1 -0.087944 0.091529 -0.961<br>Sterne_rep2 -0.255536 0.090998 -2.808<br>Sterne_rep3 -0.313482 0.078498 -3.994<br>Sterne_rep4 -0.217156 0.076043 -2.856<br>Sterne_rep5 -0.001289 0.075911 -0.017<br>timepointpost:meal_cat_corr_2021VEG*N 0.324383 0.140662 2.306<br>timepointpost:Sterne_rep1 0.819716 0.116346 7.046<br>timepointpost:Sterne_rep2 0.345521 0.115637 2.988<br>timepointpost:Sterne_rep3 -0.004239 0.099709 -0.043<br>timepointpost:Sterne_rep4 -0.233999 0.096631 -2.422<br>timepointpost:Sterne_rep5 -0.411463 0.096485 -4.265<br>meal_cat_corr_2021VEG*N:Sterne_rep1 0.013247 0.134860 0.098<br>meal_cat_corr_2021VEG*N:Sterne_rep2 0.033552 0.132654 0.253<br>meal_cat_corr_2021VEG*N:Sterne_rep3 0.113656 0.117769 0.965<br>meal_cat_corr_2021VEG*N:Sterne_rep4 0.141104 0.114431 1.233<br>meal_cat_corr_2021VEG*N:Sterne_rep5 0.076491 0.114298 0.669<br>timepointpost:meal_cat_corr_2021VEG*N:Sterne_rep1 -0.581310 0.171577 -3.388<br>timepointpost:meal_cat_corr_2021VEG*N:Sterne_rep2 -0.462081 0.168816 -2.737<br>timepointpost:meal_cat_corr_2021VEG*N:Sterne_rep3 -0.332452 0.149848 -2.219<br>timepointpost:meal_cat_corr_2021VEG*N:Sterne_rep4 -0.333598 0.145614 -2.291<br>timepointpost:meal_cat_corr_2021VEG*N:Sterne_rep5 -0.260018 0.145473 -1.787<br><br>post-meal*plant-based*5stars: b = -0.26, t = -1.8<br><br>**$p < 2.2 \times 10^{-5}$** | | Fixed effects:<br>Estimate Std. Error t value<br>(Intercept) 3.31829 0.06417 51.710<br>timepointpost 0.18425 0.06933 2.657<br>meal_cat_corr_2021VEG*N 0.15031 0.09509 1.581<br>Sterne_rep1 0.03156 0.07870 0.401<br>Sterne_rep2 -0.05081 0.07825 -0.649<br>Sterne_rep3 0.07212 0.06751 1.068<br>Sterne_rep4 0.17088 0.06539 2.613<br>Sterne_rep5 0.23545 0.06527 3.607<br>timepointpost:meal_cat_corr_2021VEG*N -0.11059 0.10424 -1.061<br>timepointpost:Sterne_rep1 -1.42087 0.08643 -16.439<br>timepointpost:Sterne_rep2 -0.69272 0.08579 -8.074<br>timepointpost:Sterne_rep3 -0.19459 0.07404 -2.628<br>timepointpost:Sterne_rep4 0.14746 0.07177 2.055<br>timepointpost:Sterne_rep5 0.42873 0.07166 5.983<br>meal_cat_corr_2021VEG*N:Sterne_rep1 -0.07957 0.11592 -0.686<br>meal_cat_corr_2021VEG*N:Sterne_rep2 -0.04480 0.11402 -0.393<br>meal_cat_corr_2021VEG*N:Sterne_rep3 -0.12283 0.10123 -1.213<br>meal_cat_corr_2021VEG*N:Sterne_rep4 -0.08433 0.09835 -0.857<br>meal_cat_corr_2021VEG*N:Sterne_rep5 -0.11195 0.09823 -1.140<br>timepointpost:meal_cat_corr_2021VEG*N:Sterne_rep1 0.34844 0.12723 2.739<br>timepointpost:meal_cat_corr_2021VEG*N:Sterne_rep2 0.01066 0.12502 0.085<br>timepointpost:meal_cat_corr_2021VEG*N:Sterne_rep3 0.13271 0.11102 1.195<br>timepointpost:meal_cat_corr_2021VEG*N:Sterne_rep4 0.07308 0.10790 0.677<br>timepointpost:meal_cat_corr_2021VEG*N:Sterne_rep5 0.05756 0.10779 0.534<br><br>post-meal*plant-based*5stars: b = 0.06, t = 0.5<br><br>**$p = .0025$** | |

*Significant effects between linear mixed models model comparisons are marked in bold. P-values represent ANOVA model comparison of linear mixed models with fixed and random effects.*

Supplementary Table 3: Interaction effects of meal category on hunger and mood for subgroups by gender (app study only).

| | **hunger** | | **mood** | |
|---|---|---|---|---|
| **app-based (5-point)** | **animal-based** | **plant-based** | **animal-based** | **plant-based** |
| **male (n = 8291)** | | | | |
| interaction effect tp*meal_cat | post-meal*plant-based: b = 0.07, t = 2.2 **p = .031** | | post-meal*plant-based: b = -0.07, t = -2.8 **p = .005** | |
| main effect meal_cat | plant-based: b = -0.01, t = -0.6 p <.53 | | plant-based: b < 0.01, t = 0.05 p = .96 | |
| **female (n = 7418)** | | | | |
| interaction effect tp*meal_cat | post-meal*plant-based: b = 0.09, t = 3.0 **p = .003** | | post-meal*plant-based: b = -0.1, t = -4.1 **p = $4.1x10^{-5}$** | |
| main effect meal_cat | plant-based: b = -0.07, t = -3.6 **p = .0004** | | plant-based: b = 0.02, t = 0.9 p = .38 | |
| **diverse (n = 279)** | | | | |
| interaction effect tp*meal_cat | post-meal*plant-based: b = -0.08, t = -0.4 p = .70 | | post-meal*plant-based: b = 0.35, t = 1.8 p = .08 | |
| main effect meal_cat | plant-based: b = -0.2, t = -1.6 p = .12 | | plant-based: b = 0.01, t = 0.04 p = .96 | |

*Significant effects between linear mixed models model comparisons are marked in bold. P-values represent ANOVA model comparison of linear mixed models with fixed and random effects.*

Supplementary Table 4: Interaction effects of meal category on hunger and mood for subgroups according to dietary adherence (app study only).

| | **hunger** | | **mood** | |
|---|---|---|---|---|
| **app-based (5-point) n = 16135 valid** | **animal-based** | **plant-based** | **animal-based** | **plant-based** |
| **Predominantly omnivorous only (n = 11600)** | | | | |
| interaction effect tp*meal_cat | post-meal*plant-based: b = 0.109, t = 3.8 **p = .0001** | | post-meal*plant-based: b = -0.10, t = -4.4 **p = .0000114** | |
| double interaction effect tp*taste | post-meal*5stars: b = -0.38, t = -4.5 **p < 2.2x10$^{-16}$** | | post-meal*5stars: b = 0.45, t = 7.3 **p < 2.2x10$^{-16}$** | |
| triple interaction effect tp*meal_cat*taste | post-meal*plant-based*5stars: b = -0.15, t = -0.8 p = .71 | | post-meal*plant-based*5stars: b = 0.18, t = 1.4 **p = .035** | |
| **Predominantly vegetarian only (n = 3456)** | | | | |
| interaction effect tp*meal_cat | post-meal*plant-based: b = -0.03, t = -0.4 p = .67 | | post-meal*plant-based: b = -0.13, t = -2.9 **p = .004** | |
| double interaction effect tp*taste | post-meal*5stars: b = -0.74, t = -4.5 **p < 5.1x10$^{-15}$** | | post-meal*5stars: b = 0.47, t = 3.9 **p < 2.2x10$^{-16}$** | |
| triple interaction effect tp*meal_cat*taste | post-meal*plant-based*5stars: b = 0.33, t = 0.8 **p = .005** | | post-meal*plant-based*5stars: b = -0.72, t = -2.5 **p = .012** | |
| **Predominantly vegan only (n = 911)** | | | | |
| interaction effect tp*meal_cat | post-meal*plant-based: b = -0.96, t = -6.6 **p < 8.3x10$^{-11}$** | | post-meal*plant-based: b = 0.54, t = 4.5 **p < 6.99x10$^{-6}$** | |
| double interaction effect tp*taste | post-meal*5stars: b = -1.44, t = -4.5 **p < 4.2x10$^{-13}$** | | post-meal*5stars: b = 0.34, t = 1.4 **p < 2.2x10$^{-16}$** | |

| | | |
|---|---|---|
| triple interaction effect tp*meal_cat*taste | post-meal*plant-based*5stars: b = -1.4, t = -1.8<br><br>p = .077 | post-meal*plant-based*5stars: b = 0.06, t = 0.11<br><br>**p = .045** |

*Significant effects between linear mixed models model comparisons are marked in bold. P-values represent ANOVA model comparison of linear mixed models with fixed and random effects.*
